# Prospective metagenomic sequencing of wastewater across the United States yields robust viral enrichment and concordance with digital PCR measurements

**DOI:** 10.64898/2026.05.07.26352651

**Authors:** Marlene K. Wolfe, Devin North, Alexander L. Jaffe, Alessandro Zulli, Dorothea Duong, Bridgette Shelden, Miriam Goldman, Miles Richardson, Peter Thana, Vikram Chan-Herur, Pouya Kheradpour, Amanda L. Bidwell, Stephen P. Hilton, Sheena Conforti, Abigail P. Paulos, Alexandria B. Boehm

## Abstract

Metagenomic sequencing is increasingly applied to wastewater to characterize the diversity, dynamics, and relative abundance of human and animal viruses. Among these sequencing approaches are those that enrich viral nucleic acids from the wastewater matrix, aiming to increase the viral read fraction for analysis. However, the feasibility of scaling targeted viral sequencing to diverse sewersheds across large geographic scales is currently unknown. In this study, we apply hybrid capture metagenomic sequencing to nearly 450 weekly wastewater samples collected during the respiratory virus season in the United States and evaluate sequencing performance for generating public health-relevant data. Analysis of data from 15 wastewater treatment plants demonstrates that our approach enabled efficient capture of pathogens of interest, achieving a median viral read fraction over 19%. Importantly, relative abundance estimates of common pathogens correlated with direct quantification of viral targets using RT-ddPCR. Together, our results demonstrate that hybrid capture sequencing of wastewater is a viable tool to monitor both common and rare pathogens across geographically diverse sewersheds.

## Introduction

Wastewater-based epidemiology (WBE) is a valuable public health tool for monitoring the occurrence of viral infectious diseases including COVID-19, influenza, mpox, and measles^1–4^. Public health agencies, including local, regional, and national programs, consider WBE an important and actionable source of epidemiological information, informing early outbreak detection and guiding public health responses^5,6^. As such, a large number of efforts globally measure viral biomarkers prospectively in wastewater^7^. Prospective monitoring efforts largely use polymerase chain reaction (PCR) to detect viral genomes in wastewater. Quantitative formats such as qPCR or digital droplet PCR offer measurement precision and rapid turnaround times^8^. However, PCR methods have several key limitations, including the limited number of different assays that can be run together and the need for highly specific and sensitive oligo primers and probes. As viral genomes can evolve over very short time scales^9^, primer and probe sequences may require frequent adjustment to ensure assays remain sensitive and specific to circulating variants^10^. Primers and probes can only be designed if a viral genomic sequence is known, limiting PCR assay use for highly novel viruses or divergent variants of known viruses.

Genomic surveillance via sequencing offers an attractive alternative to quantitative PCR approaches for viral WBE^11^. Tiled amplicon sequencing to characterize SARS-CoV-2 variants has been widely used for years to understand viral evolution, applied to both clinical^12^ and wastewater samples^13^. Application to wastewater samples has demonstrated close concordance with clinical results when analyzing SARS-CoV-2 lineages^12^ and can provide important information on the evolution of SARS-CoV-2 in areas without clinical testing^13^. Tiled amplicon approaches have also been applied to sequencing influenza^14^, enterovirus D68, norovirus GII, human adenovirus 41, hepatitis A virus, and measles^15^ genomes in wastewater. A major drawback to tiled amplicon approaches is that they focus on a single virus.

Recently, several studies have applied metagenomic approaches to viral pathogen WBE^15,16^. Since viral nucleic acids represent a small fraction of the total nucleic acids in wastewater^17^, shotgun metagenomic approaches – which generally employ few, if any, steps to enrich for targets of interest – can fail to detect lower-abundance viruses, resulting in highly variable estimates of sensitivity^18^. There are two approaches to overcome this limitation. The first is to sequence deeply, generating billions of sequencing reads per sample, thereby enabling the detection of rare viral targets^16,19^. However, at present, deep, untargeted sequencing may not be feasible for prospective viral WBE efforts due to cost and may not provide the high viral genomic coverage needed to identify the emergence of new variants. The second approach uses probe-based hybrid capture to enrich for viral targets before sequencing^20,21^. This ‘semi-targeted’ approach promises to enrich viral sequencing reads by orders of magnitude over shotgun sequencing approaches while being tolerant to up to 30% sequence divergence from known references^22,23^. Additionally, previous work conducted at a single WWTP in New Haven, CT, USA^24^ found viral sequence read counts obtained using a hybrid capture sequencing correlated positively with digital PCR measurements of respiratory and enteric viral nucleic acids. However, the performance of hybrid capture approaches relative to targeted molecular methods under varying sampling conditions remains poorly characterized.

In this study, we evaluate the feasibility of hybrid capture metagenomic sequencing to detect viral biomarkers in wastewater samples collected weekly from 15 wastewater treatment plants (WWTPs) across the U.S. We examine factors impacting the performance of our sequencing method, including the extent of viral diversity recovered, and compare the relative abundance of sequenced viruses to concentrations measured by reverse-transcriptase droplet digital PCR (RT-ddPCR). Spatio-temporal dynamics across the study are presented for select viruses.

## Methods

Wastewater samples were collected and processed to obtain viral metagenomic sequences and determine the concentration of viral targets using reverse transcription digital droplet PCR (RT-ddPCR). Viral nucleic acids sorb to wastewater solids and are enriched orders of magnitude on a per mass basis in wastewater solids compared to liquids^25,26^, therefore, nucleic acids obtained from wastewater solids represent good templates for sensitivity and specific ddPCR assays^8^; so, nucleic acids obtained from solids were used to measure viral biomarkers using RT-ddPCR. Wastewater solids consist of all sorts of particles larger than 0.2 µm in diameter, including bacterial and eukaryotic cells and debris, and inorganic particles. When the ratio of non-viral to viral sequences is high, as is the case in wastewater solids, probe-capture viral metagenomic sequencing methods may not adequately identify viral sequences^27^; pilot work suggests this to be the case in our laboratory. As such, probe-based metagenomic sequencing was applied to wastewater liquids, in contact with the wastewater solids, as described below.

### Sample collection and prospective measurements of viral nucleic acids in wastewater solids

Wastewater samples were collected approximately three times per week for 12 weeks from 15 wastewater treatment plants (WWTPs) (n=443 samples) across 12 states in the United States (Fig. 1, Table 1). WWTP locations were chosen to represent a variety of geographic regions and communities, with WWTPs serving populations ranging from 31,000 to 3.5 million across the Midwest, Northeast, South, and West regions of the US. Sample collection used sterile technique and followed the protocol outlined in Boehm et al.^10^ Two sample types were collected depending on location and based on WWTP staff preference: grab samples of sludge from the primary clarifier or 24-hour composite primary influent samples (Fig. 1, Table 1). Samples were shipped overnight at 4°C to the laboratory for processing.

**Figure 1.**
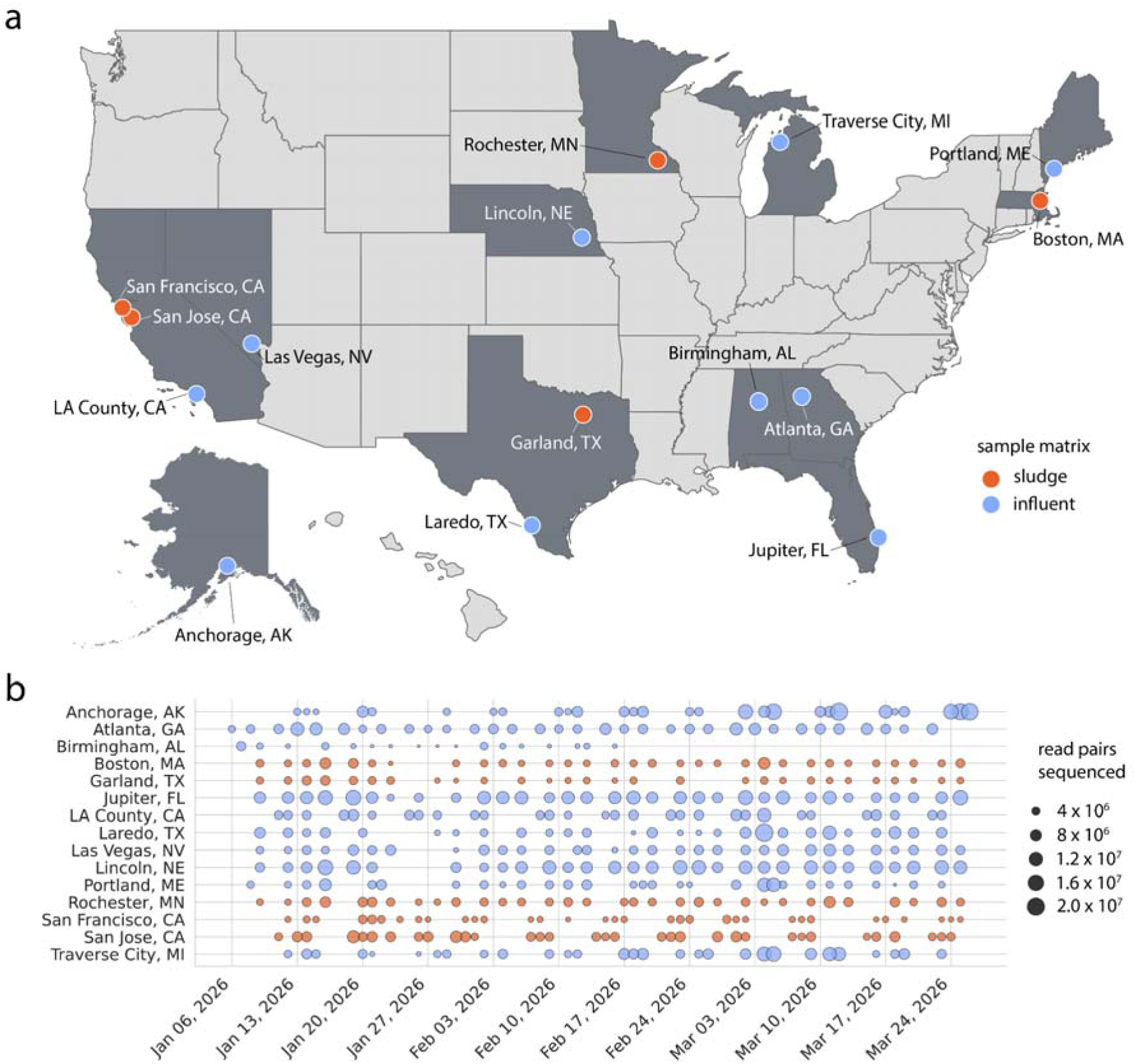
Characteristics of samples analyzed in this study. **a)** Map of the 15 wastewater treatment plants (WWTPs) enrolled in this study, along with sample type. States with participating WWTPs are shaded in dark gray. Alaska and Hawaii are not to scale. **b)** Temporal frequency of sampling, with the size of points indicating the total number of read pairs sequenced after human read filtering and de-duplication.

**Table 1.** Characteristics of sites included in the study, including location, population served, sample matrix, total number of samples collected per site, and sampling dates (month/day/year format).

| Site | Location | Population served | Sample Matrix | Total # samples | Start date | End date |
| --- | --- | --- | --- | --- | --- | --- |
| John M. Asplund Water Pollution Control Facility | Anchorage, Alaska | 220,000 | Primary Influent | 28 | 1/13/26 | 3/26/26 |
| RM Clayton Water Reclamation Center | Atlanta, Georgia | 600,000 | Primary Influent | 33 | 1/6/26 | 3/22/26 |
| Village Creek Water Reclamation Facility | Birmingham, Alabama | 200,000 | Primary Influent | 17 | 1/7/26 | 2/16/26 |
| Deer Island Treatment Plant | Boston, Massachusetts | 2,400,000 | Sludge | 31 | 1/9/26 | 3/25/26 |
| City of Garland Rowlett Creek Wastewater Treatment Plant | Garland, Texas | 630,000 | Sludge | 28 | 1/9/26 | 3/25/26 |
| Loxahatchee River Environmental Control District | Jupiter, Florida | 90,000 | Primary Influent | 32 | 1/9/26 | 3/25/26 |
| A.K. Warren Water Resource Recovery Facility | Los Angeles County, California | 3,500,000 | Primary Influent | 30 | 1/11/26 | 3/23/26 |
| Zacate Creek Wastewater Treatment Plant | Laredo, Texas | 140,000 | Primary Influent | 27 | 1/9/26 | 3/23/26 |
| Clark County Water Reclamation District Flamingo Water Resource Center | Las Vegas, Nevada | 1,250,000 | Primary Influent | 31 | 1/9/26 | 3/25/26 |
| Theresa Street Water Resource Recovery Facility | Lincoln, Nebraska | 240,000 | Primary Influent | 30 | 1/9/26 | 1/9/26 |
| Portland Water District East End Wastewater Treatment Facility | Portland, Maine | 65,000 | Primary Influent | 29 | 1/8/26 | 3/23/26 |
| City Of Rochester MN Water Reclamation Plant | Rochester, Minnesota | 120,000 | Sludge | 32 | 1/9/26 | 3/25/26 |
| Southeast San Francisco Wastewater Treatment Plant | San Francisco, California | 750,000 | Sludge | 33 | 1/12/26 | 3/25/26 |
| San Jose-Santa Clara Regional Wastewater Facility | San Jose, California | 1,500,000 | Sludge | 33 | 1/11/26 | 3/24/26 |
| Traverse City Regional Waste Water Treatment Plant | Traverse City, Michigan | 30,623 | Primary Influent | 29 | 1/12/26 | 3/23/26 |

Immediately upon receipt, aliquots from samples were centrifuged, and the pellets, representing wastewater solids, were processed for viral nucleic acid quantification (Fig. 2). Targets included nucleic acids of SARS-CoV-2, influenza A and B, respiratory syncytial virus, human metapneumovirus, enterovirus D68, measles, norovirus GII, rotavirus, hepatitis A, parvovirus, and West Nile virus. Quantification was performed using the methods described in detail elsewhere^8^; the exception is that on 2 March 2026, a second measles probe was added to account for sequence divergence observed in some circulating genotypes (see SI). Concentrations are reported as copies per gram of dry-weight solids.

**Figure 2.**
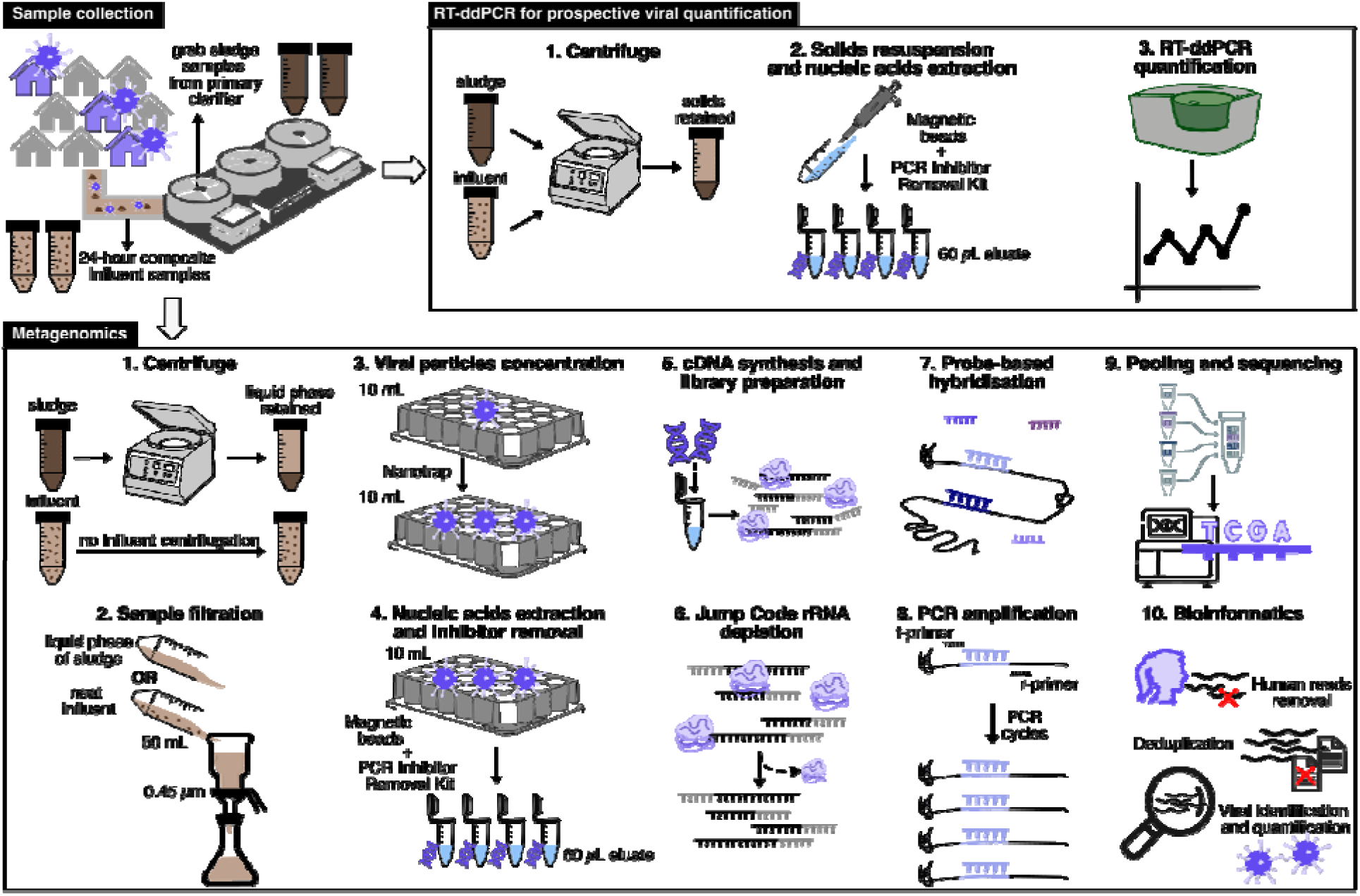
Workflow of wastewater samples processing and comparison between RT-ddPCR and metagenomics sequencing approaches. Abbreviations: RT-ddPCR, reverse transcription digital droplet Polymerase Chain Reaction; cDNA, complementary DNA; rRNA, ribosomal RNA.

### Pre-analytical processing, nucleic acid extraction, and purification for metagenomic sequencing

Pre-analytical processing and nucleic acid extraction and purification for metagenomic sequencing were aimed at minimizing the presence of inhibitors and the ratio of non-viral (bacterial and eukaryotic) to viral targets^27^ prior to sequencing. Sludge samples from the primary clarifier were centrifuged at 4200×g for 5 minutes, and 50 mL of the supernatant was collected. Fifty mL primary influent samples were processed without centrifugation. The 50 mL aliquots were stored between 0 and 8 days at 4°C and processed in weekly batches.

Sludge supernatants and influent samples were vacuum-filtered using 0.45 μm pore-size polyethersulfone (PES) filters (Thermo Scientific #165-0045, Waltham, Massachusetts, USA) to remove bacterial and eukaryotic nucleic acids, and the filtrates were collected in sterile containers. Viral particles were then concentrated from 10 mL of filtrate using Ceres Nanotrap Microbiome A particles (Manassas, Virginia, USA) following manufacturer’s directions.

Nucleic acids were extracted from the concentrated viral particle fraction using the MagMAX Viral/Pathogen Nucleic Acid Isolation Kit (Waltham, Massachusetts, USA) using an automated KingFisher instrument (ThermoFisher Scientific, Waltham, MA, USA). PCR inhibitors were removed using the Zymo OneStep PCR Inhibitor Removal Kit (Irvine, California, USA). The final volume of the nucleic acid extract obtained from 10 mL of filtrate was 60 µl.

### Library preparation and sequencing

cDNA synthesis was performed with ProtoScript II First Strand cDNA Synthesis Kit (New England Biolabs #E6560L, Ipswich, MA, USA) using the neat nucleic acid as template. cDNA was purified using Kapa HyperPure Beads (Roche, Basel, Switzerland), and the cDNA concentration was measured using Qubit (Thermo Fisher). cDNA was then diluted to 1 ng/μL using nuclease-free water for library preparation. The Twist Library Preparation Kit (Twist Biosciences, South San Francisco, CA, #103548) was used for enzymatic fragmentation, adapter ligation, and PCR amplification, with the addition of the Twist Unique Molecular Identifiers (UMI) adapter system.

Prepared libraries were enriched using the Twist Comprehensive Viral Research Panel (Twist Biosciences, #103548), which, as reported by the manufacturer, targets more than 3000 viral genomes, through hybridization at 70°C for 16 hours. Bacterial, human, and other mammalian ribosomal RNA (rRNA) was depleted from the samples using CRISPR-Cas9 technology via the CRISPRclean Stranded Total RNA Prep with rRNA Depletion kit (Jumpcode Genomics, San Diego, CA #KIT1014). Prepared libraries were sequenced on an Illumina (Foster City, CA) NovaSeq X Plus (10B flow cells, 150-bp paired-end reads), alongside negative sequencing controls. Negative sequencing controls used DNA/RNA-free water as template for the cDNA synthesis and were moved through the workflow as samples for each plate (11 total were processed). Samples were pooled in batches of 8, with a target sequencing depth of 100 million reads per sample. Full library preparation details are provided on protocols.io^28^.

### Computational analyses

After demultiplexing, reads were deduplicated using HUMID (https://github.com/jfjlaros/HUMID), incorporating information from the unique molecular identifiers (UMIs). We next performed host read removal by iteratively applying Kraken2 (v2.1.3), bowtie2 (v2.5.2) with reference GRCh38.p14, and HRRT (v2.2.1) with the human_filter.db.20250916v2 database. Additionally, raw sequencing reads were trimmed of sequencing adapters and UMIs using fastp (v0.23.4). Taxonomic classification and read quantification were computed using EsViritu^29^ (v1.1.0) with database version 3.2.2. This approach employs competitive alignment with minimap2 against a curated database of high-quality virus genome assemblies, enabling simultaneous detection of many viruses. Intermediate alignment files were retained (*--keep True)*, and the initial, unfiltered BAM file was used to compute the number of putative viral read pairs for each sample (Table S1).

Specifically, read pairs were designated as viral if one or both mates aligned to an Esviritu reference sequence during the initial mapping. Putative rRNA reads were classified using sortmerrna^30^ (v4.3.6) with the smr_v4.3_sensitive_db.fasta database and an alignment e-value threshold of 1*10^-5^. Read pairs that were preliminarily classified as both viral and rRNA (typically <0.1%) were designated as ambiguous and removed from final viral and rRNA read counts (Table S1).

To examine off-target sequences generated by our protocol, we filtered out reads initially classified as viral or as rRNA using the tools above, and assembled the remaining reads using two variants of the SPAdes assembler (<u>metaspades.py</u> and <u>rnaspades.py</u>)^31^. Resulting contigs were filtered to those at least 1000 bp in length. Focusing on three sites – Las Vegas, Traverse City, and Atlanta – we combined all filtered contigs on a per-site basis and clustered them using mmseqs (*easy-linclust --min-seq-id 0.95 --cluster-mode 2 --cov-mode 1*), reasoning that there would be significant sequence redundancy from high-frequency sampling of the same sewersheds over time. For each site, raw metagenomic reads for each sample were competitively mapped back to their corresponding contig set using bowtie2^32^ and samtools^33^. Alignment files were processed using CoverM^34^ with both loose (--min-read-percent-identity 0) and stringent (--min-read-percent-identity 0.95) mapping parameters, generating per-contig read counts and coverage breadth values for downstream analyses. Contigs were considered ‘detected’ in a sample if their breadth of coverage at 95% read identity met or exceeded 50%. Preliminary classifications were made for contigs recruiting 50,000 or more reads in any sample using BLAST against core_nt and refseq_rna (blastn).

### Direct validation

For a subset of targets classified as associated with high-consequence diseases (HCD), manual confirmation of classification was performed. HCDs are associated with serious illness with high case fatality rates. The list of high-consequence pathogens for this study (Table S2) was based on global HCD classifications or on pathogens that share high genetic similarity with these pathogens^35^. For a flagged target, reads were manually inspected using BAM files from the EsViritu output. NCBI BLAST was used to manually classify the reads against the entirety of NCBI’s core_nt database using the blastn algorithm^36,37^. Exclusionary BLAST was also performed to ensure there were no other potential classifications. Lastly, sequences were BLASTed against the NCBI patent database to identify potential synthetic matches. If the EsViritu-based classification was confirmed, further analysis of the sequences was performed by comparing them to known synthetic vectors and determining whether multiple classified reads were distributed across the flagged target genome. For reads classified as poliovirus, a pipeline was built to BLAST them against a curated database of complete poliovirus genomes (polio-blast-validation v.0)^38^. Alignment of the reads was also performed to determine which genomic region they were present in. Reads were then manually inspected as described above, with only reads in the protein capsid region considered informative.

### Statistical analyses

Sequence run quality was assessed using the duplication rate, rRNA sequence relative abundance, and the richness and diversity of viral species. Duplication rate was defined as the percentage of total sequencing reads that were redundant after UMI/sequence-based clustering by HUMID (see above). rRNA relative abundance was defined as the fraction of total reads mapped to ribosomal RNA sequences. The number of unique viral species (viral richness) and the Shannon diversity index were calculated using the vegan R package^39^. We compared alpha diversity over time for each site using a linear regression. A Bray-Curtis dissimilarity matrix was used to compare viral species composition, defined as relative abundance (number of detected reads of a specific target divided by the total number of filtered reads in the sample), among all samples. Then, non-metric multidimensional scaling (NMDS) ordinations were used to visualize results, with sample placement in the NMDS ordination colored according to WWTP and sample matrix. A PERMANOVA was used to test the null hypothesis of no difference in viral community composition between groupings (WWTP and sample matrix, tested separately). This beta diversity analysis was also performed using the vegan R package^39^.

Results from hybrid capture sequencing and RT-ddPCR results were compared for a short list of viral targets: SARS-CoV-2, influenza A, influenza B, RSV (inclusive of subtypes A and B), human metapneumovirus (HMPV), rotavirus A, norovirus GII, adenovirus F, measles, hepatitis A, and West Nile virus. RT-ddPCR results were compared to results for the same viral species based on taxonomic classification and read quantification from EsViritu; the list of species used for each viral target is listed in Table S3. The exception is Norovirus GII, where only GII subspecies were retained for comparison. We evaluated both agreement in detection across samples and the association between the abundance estimates for each sample. Agreement between hybrid capture sequencing and RT-ddPCR results was evaluated using Percent Positive Agreement (PPA), Percent Negative Agreement (PNA), and Overall Percent Agreement (OPA). PPA is the percentage of samples that tested positive for a given target by both sequencing and RT-ddPCR, and PNA is the percentage of samples that tested negative by both methods. OPA is the overall percentage of samples for which RT-ddPCR and sequencing results were concordant, either both negative or both positive. PPA, PNA, and OPA were computed across all sites and for each site individually for every viral target measured using both methods. To evaluate the overall association between RPKMF and gc/g while controlling for city and virus effects, as well as repeated-measures testing for viruses within each sample, a linear mixed model was used. Measurements below the limit of detection were replaced with values approximately half the estimated limit of detection (0.001 for RPKMF and 500 for gc/g), and all measurements were log-transformed prior to analysis with the lme4 package in R. Random intercepts were included for city, virus, and city/date to account for these differences and repeated measures. The performance package in R was used to compute conditional and marginal R^2^. When a viral target was detected by both methods, Kendall’s tau was used to assess the association between sequencing reads (measured in reads per kilobase of genome per million filtered reads (RPKMF), with filtered reads referring to reads passing initial quality control) and RT-ddPCR results at each site. For Kendall’s tau, p-values were adjusted for multiple comparisons using the Benjamini-Hochberg false discovery rate correction, but no adjustment was made for repeated measures.

## Results

### Assessing sequencing quality of hybrid capture metagenomes

After human read removal and deduplication, a median of 4.9*10^6^ unique read pairs were sequenced across the 443 samples (Fig. 1b), with a median duplication rate of 0.97 (IQR 0.97 - 0.98) (Table S1, S4). Deduplicated, de-humanized reads were then subjected to viral classification, revealing a median of 8.4*10^5^ viral read pairs (19.4%) across the 443 samples, with substantial variation across sites (Fig. 3a, Table S1). Notably, samples collected from five sites – Jupiter, Lincoln, Traverse City, Las Vegas, and Anchorage – attained median viral read percentages over 25% (N = 32, 30, 29, 31, and 28 for the sites, respectively) (Fig. 3b).

**Figure 3.**
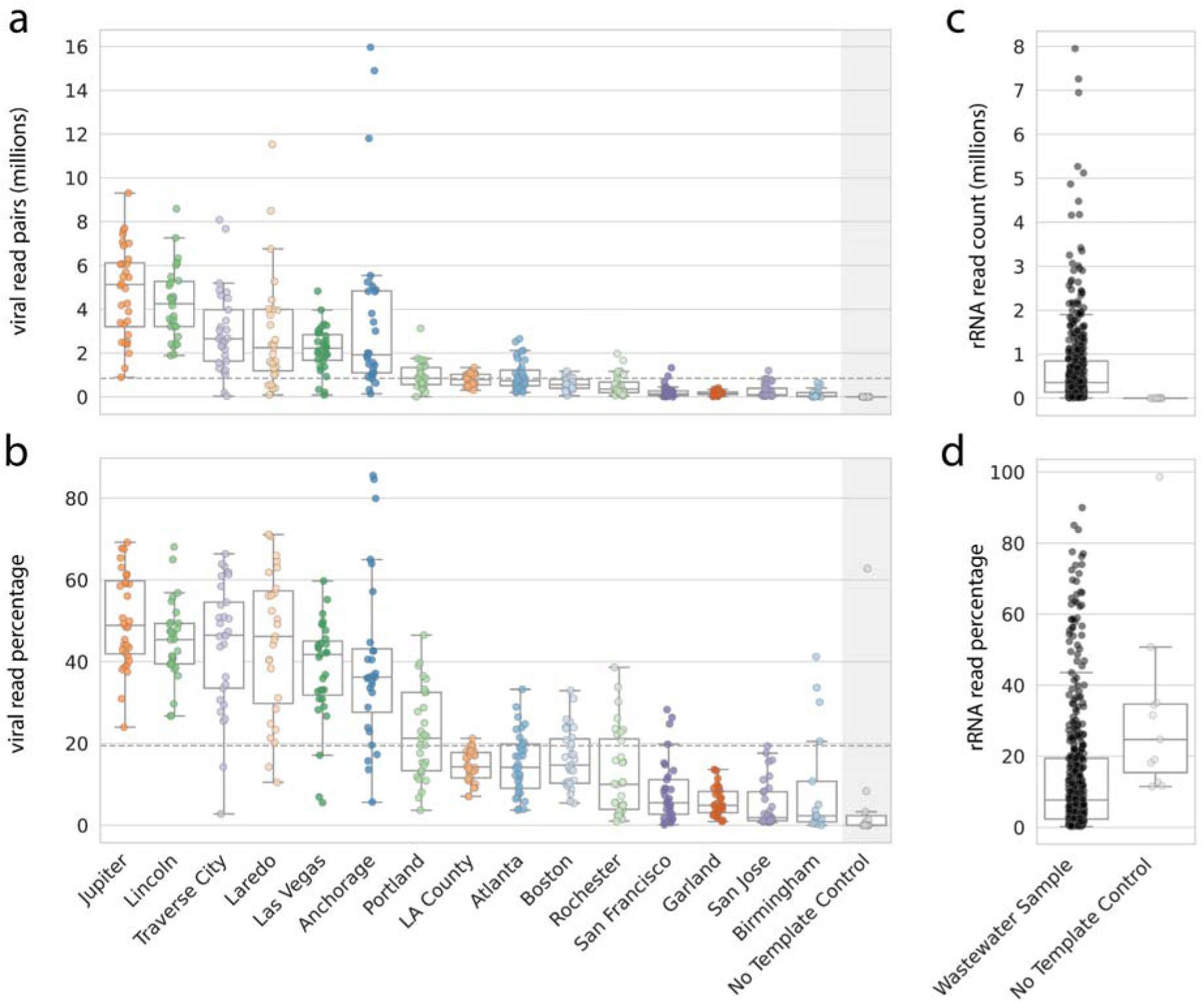
Components of hybrid capture metagenomes. **a)** Viral read pairs classified per site and **b)** percentage of total, deduplicated read pairs classified as viral. Dashed lines indicate the median values across all samples, excluding no template controls. **c)** Number and **d)** percentage of total, deduplicated read pairs classified as rRNA by sample category.

We investigated the relationship between the total number of sequenced read pairs and the number of viral read pairs recovered, observing a positive, linear relationship between log-transformed values (Fig. S1). This positive, linear trend was particularly apparent for influent-derived samples (R^2^ = ∼0.73). Similar trends were observed between the total number of read pairs sequenced and the number of unique, high-confidence viral detections per sample (Methods) after log transformation (R^2^ = ∼0.68). rRNA reads were relatively abundant in sequenced read sets (median ∼3.6*10^5^ / ∼7.7%) despite efforts to reduce off-target nucleic acids during processing (Fig. 3cd, Table S1).

Eleven no-template controls (NTCs) were processed to evaluate the potential for reagent and/or other sources of contamination during library preparation. Very few viral reads were sequenced in these controls (fewer than 300 viral read pairs per sample across all controls), and rRNA made up a higher proportion of reads in NTCs compared to wastewater samples (Table S5, Fig. 3c, 3d). However, in the cases where viral reads were observed (6/11 NTCs), they were predicted to derive from only four viruses - human mastadenovirus F, mamastrovirus 1, human papillomavirus 28, and Acheta domestica mini ambidensovirus – with read counts generally far below those observed in wastewater samples (Fig. S2). Given this, we inferred that widespread reagent contamination is unlikely for these samples, though we cannot entirely rule out the possibility of some sporadic well-to-well contamination, a common phenomenon in low-biomass metagenomic studies^40^.

Sequencing runs were considered to be high-quality if they did not exceed a 99% duplication rate and produced ≥10^6^ unique reads. These thresholds were chosen as they appeared to separate samples with high and low numbers of viral read pairs (Fig. S1). Overall, 94.6% of samples (419/443) were considered high quality (420/443 with <=99% duplication rate, 437/443 with >=10^6^ unique reads). Performance varied by site, with the percentage of high-quality samples at an individual site ranging from 29.4% (Birmingham) to 100% at 9 of the 15 sites (Table S4). One sample from Birmingham yielded no viral detections (1/26/2026). Sequencing performance, measured as the number of viral reads and the duplication rate, was compared between samples derived from influent wastewater and those derived from sludge. The number of viral read pairs obtained using influent was larger than the number obtained using sludge (difference in median viral read pairs = 4.9*10^6^, Kruskal-Wallis H = 85.8, p = 2.0*10^-20^, n = 443). There was no difference in the duplication rate between the two sample types (Fig. S3). Birmingham sample sequencing was terminated after 17 samples on Feb 16 (Table S1) because samples from the site consistently failed to meet quality thresholds and showed low levels of detection of common viruses such as norovirus.

### Sample composition of hybrid capture metagenomes

From the ∼760 million viral read pairs obtained from 443 wastewater samples, 1,334 unique viral species and 2,227 viral subspecies were identified across 50 viral families. Across all samples, the most abundant viral families were typically *Sedoreoviridae* (rotaviruses) and *Astroviridae* (astroviruses), which together accounted for over 75% of reads in most samples. *Caliciviridae* (noroviruses) and *Adenoviridae* (adenoviruses) were the next most abundant families, accounting, respectively, for 2.0-19.7% and 1.3-55.1% of reads on average (Fig. S4a.). In most locations, *Adenoviridae* (adenoviruses) accounted for ∼2-8% of reads across all samples; however, in certain locations (Rochester, San Francisco, and San Jose), *Adenoviridae* was much more prevalent (∼30-50%, with some samples having >90%) (Fig. S4a.). Notably, all locations with increased relative abundances of *Adenoviridae* provided sludge samples. Other common human viral pathogen families, such as *Polyomaviridae* (polyomaviruses), *Coronaviridae* (coronaviruses), *Papillomaviridae* (HPV), and *Picobirnaviridae* (enteroviruses), were identified in over 97% (434/443) of samples, while *Paramyxoviridae* (parainfluenza) and *Orthomyxoviridae* (influenza) were present in 76% (337/443) and 83% (369/443), respectively (Fig. S4b).

Across 443 samples, a median of 230 viral species (minimum: 6, maximum: 357) was found in each sample, with the most abundant species being human adenovirus F (*Human mastadenovirus F*), human astrovirus (*Mamastrovirus hominis*), and rotavirus A (*Rotavirus alphagastroenteritidis*) found in 442/443 samples. For this analysis, we refer to viruses using their common names; the corresponding species names can be found in Table S3. Common respiratory pathogens were found in a majority of samples, concordant with the winter onset periods of many of these diseases; SARS-CoV-2, influenza A, rhinovirus A and C, and respiratory syncytial virus (RSV) were found in >71% of samples (316/443) (Fig. S5.). Human metapneumovirus (HPMV) was detected in 243/443 (55%) samples. During the study period, many other pathogens of clinical relevance were found, including Epstein-Barr virus, cytomegalovirus, measles, and chickenpox, in 69%, 42%, 6.3%, and 5.0% of samples, respectively. Multiple strains of human papillomavirus (HPV), both high- and low-risk, as well as hepatitis A, B, C, and E, were detected (Fig. S5.). Heatmaps showing the relative abundances of 75 viral targets (highlighted in table S3) at each sampling location can be found in the supplementary information.

Sample diversity was analyzed both temporally and spatially. Average species richness, the number of species detected in each sample, ranged from a minimum of 57 in Birmingham to a maximum of 302 in Lincoln, with a group median of 222 species. Alpha diversity, as measured via the Shannon index, ranged from 0.25 to 2.93 across all samples, with a median of 1.79 (Fig. S6a). While individual samples from multiple locations fell significantly above or below the average, no site differed significantly from the mean on average, indicating that even low-performing locations had comparable diversity yields. Throughout the sampling period, all locations except Jupiter, Portland, and Rochester showed statistically significant decreases in alpha diversity with respect to time from January to March (p < 0.05 for San Francisco, p < 0.001 for all others), despite no significant change in species richness in any location over that period (Fig. S7). To assess variation in viral diversity among sites, Bray-Curtis dissimilarity was used to compare relative abundances of all species across samples. The NMDS 2-D ordination has a stress of 0.11 suggesting a fair fit. The null hypothesis of no variation among samples due to grouping was rejected and indicated samples of the same type (influent vs sludge), (R^2^ = 0.14, p < 0.001) (Fig. S6b) and from the same WWTP (R^2^ = 0.44, p < 0.001) tended to be more similar to each other than to other samples (Fig. S6c).

To explore the off-target components of the sequenced viromes, we removed reads initially classified as viral or rRNA from metagenomic samples from three sites - Las Vegas, Atlanta, and Traverse City - and performed *de novo* assembly of the remainder to create larger contiguous DNA/RNA fragments. Assembled fragments recruited a median of ∼67.4% of filtered reads (Fig. S8a), and many were consistently detected across the time series at three sites. This was particularly true in Las Vegas, where ∼12,000 unique fragments were detected in 75% or more of samples (Fig. S8b). Among the fragments recruiting the most reads across samples (≥50,000 reads), we detected genetic signatures of protistan ribosomal RNA, potential phage sequences, and Picornaviridae viruses that were apparently not identified by our viral classification pipeline, possibly due to poor database representation.

### Relationship between quantitative sequencing and ddPCR measurements

The percentage of samples positive for a given virus was higher with RT-ddPCR than with sequencing for most targets, except for measles (Fig. 4a). When considering whether results from the two methods agreed on a per-sample basis, overall percent agreement (OPA) for a viral target ranged from 48.8% for HMPV to 99.8% for rotavirus, which was detected by both methods in all samples that produced viral sequences. Percent positive agreement (PPA) ranged from 9.4% for measles and 28.1% for hepatitis A to 99.5% for both adenovirus group F and norovirus GII and 99.8% for rotavirus (Fig. 4b). Percent negative agreement (PNA) ranged from 0% for norovirus GII, adenovirus group F, and rotavirus to 100% for West Nile Virus, which was never detected by either method. When percent agreements were grouped by both viral target and testing location, agreement varied by location and by viral target (Fig. 4c). OPA at individual locations varied the most for HMPV, from 9.1% in San Jose to 93.3% in Lincoln (although OPA was 100% for HMPV in Birmingham, the virus was not detected in any sample), followed by Influenza B, with an OPA of 9.4% in Jupiter and 89.7% in Traverse City. There were no significant differences in the median PPA, PNA, or OPA values for viral targets between locations providing influent samples and those providing sludge (Wilcoxon rank-sum test, p = 0.14, 0.33, and 0.27, respectively; Table S6).

**Figure 4.**
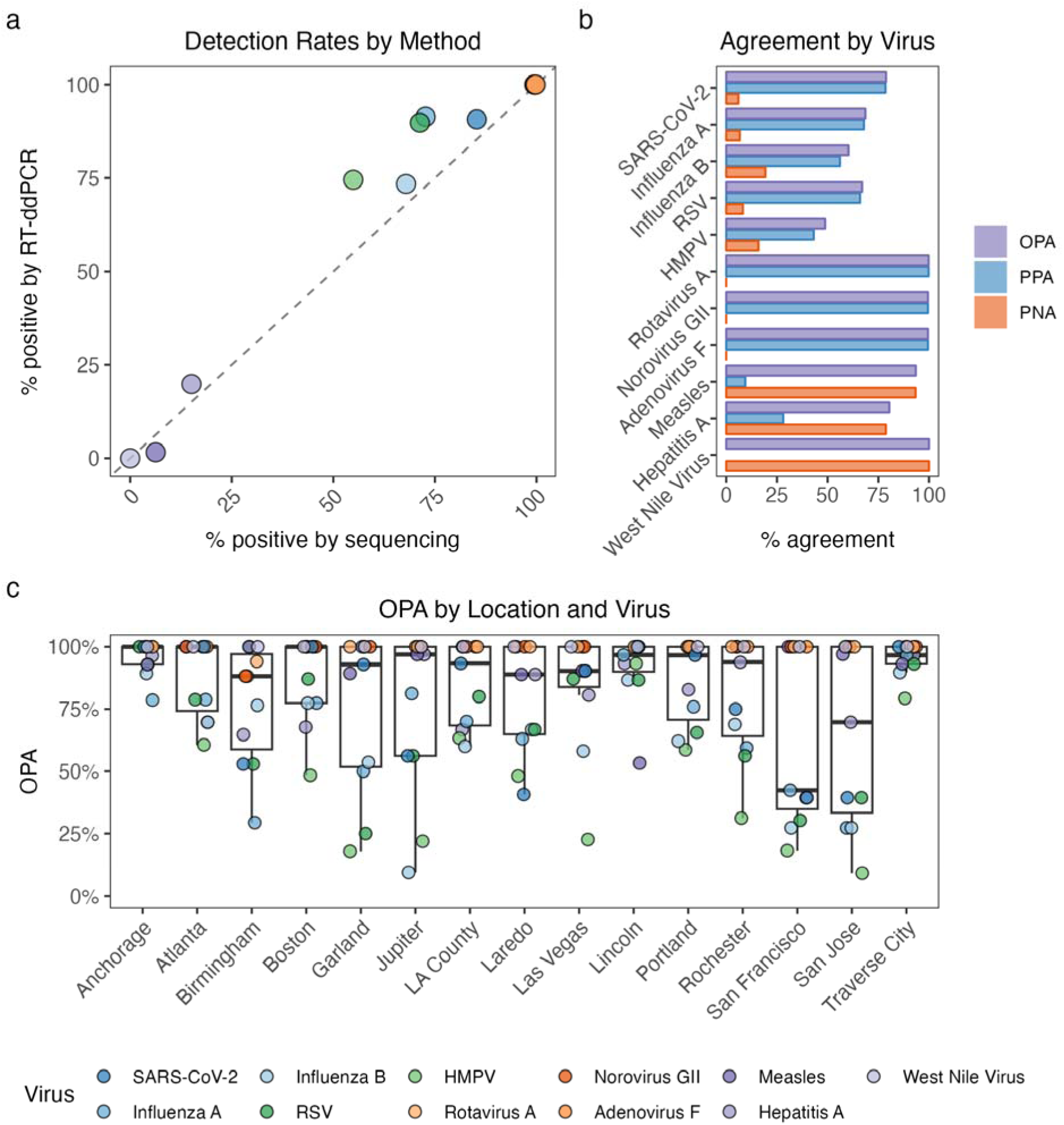
Detection rates and agreement between sequencing and RT-ddPCR. **a)** percent of samples positive by ddPCR compared to percent of samples positive by sequencing. The dashed line represents a 1:1 relationship. Norovirus, Rotavirus A, and Adenovirus F are overlapping points in the top right corner of the plot. **b)** OPA, PPA, and PNA by virus inclusive of data from all sites. **c)** OPA by sampling location (x-axis) and by viral target (color). Individual points represent OPA for each viral target at a given sampling location, and boxplots represent the distribution of OPA across the list of viral targets at each site.

A linear mixed model revealed that target concentrations measured by RT-ddPCR were a significant positive predictor of RPKMF across all measurements (slope = 0.52, 95% CI 0.47 - 0.57, t = 19.8, p < 2.0*10^16^). RT-ddPCR concentration explained 12.4% of variance (marginal R^2^), and when accounting for variation across cities and viruses and repeated sample measures, the model explained 83.5% of the total variance (conditional R^2^). The intraclass correlation coefficient (ICC) was calculated for each random effect, and the majority of total variance in the model was attributable to viral target (variance = 3.15, ICC = 71.8%) with some variation by sampling location (variance = 0.41, ICC = 9.4%). A time series of measurements at selected locations and viral targets are shown in Fig. 5, all sampling locations and viral targets are shown in Fig. S9.

**Figure 5.**
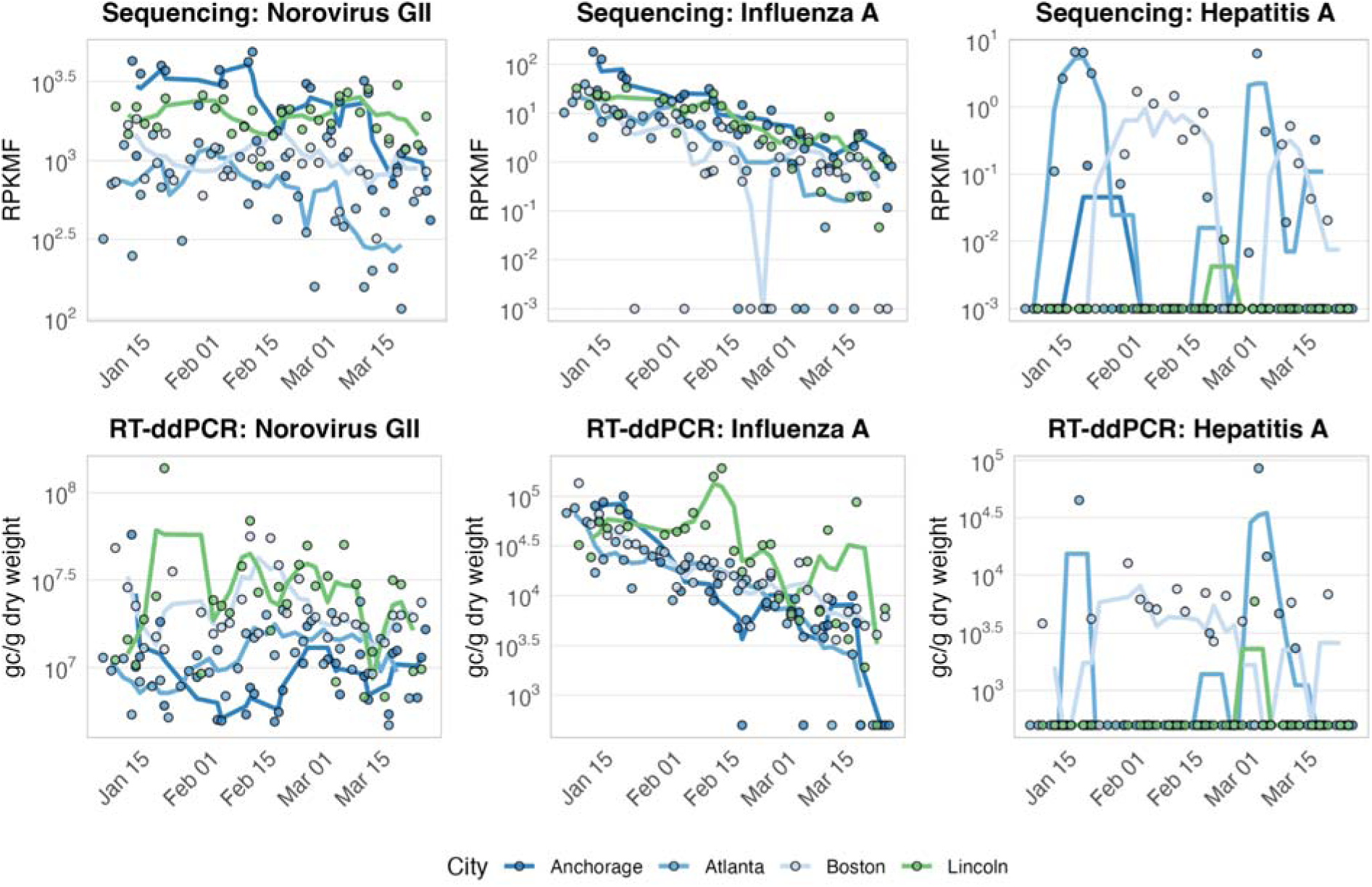
Time series of relative abundance of select viral targets (norovirus GII, influenza A, and hepatitis A) via sequencing (RPKMF, top row) and abundance in gc/g by RT-ddPCR (bottom row) for four sites, one from each region of the United States. Points represent values for individual samples, lines represent a 3-sample rolling average. A full time series for all targets is available in the SI. Note that y-axis scales are specific to each plot.

When Kendall’s tau was used to investigate associations within each sampling location for all viral targets considered together, there was a strong and significant association between RPKMF and target concentrations measured by RT-ddPCR with a median Kendall’s tau = 0.51 across all sites, ranging from tau = 0.42 in San Jose (adjusted p = 1.6*10^-22^, n = 275) to tau = 0.61 in Lincoln (adjusted p = 2.5*10^-46^, n = 256) (Fig. 6a, Table S6). There was a small but significant difference between tau for samples collected at locations providing influent compared to sludge (Wilcoxon rank-sum test, p = 0.04, median tau for influent = 0.54, median tau for sludge = 0.50). When results were grouped by viral target across all locations the association between the two methods was not as strong, with a median of tau = 0.07 across all sites (Fig. 6b). There was no significant association across sites for RSV, measles, norovirus, or hepatitis A. Among the significant associations, tau ranged from -0.14 for HMPV (adjusted p = 1.3*10^-4^) to tau = 0.27 for SARS-CoV-2 (adjusted p = 2.9*10^-16^).

**Figure 6.**
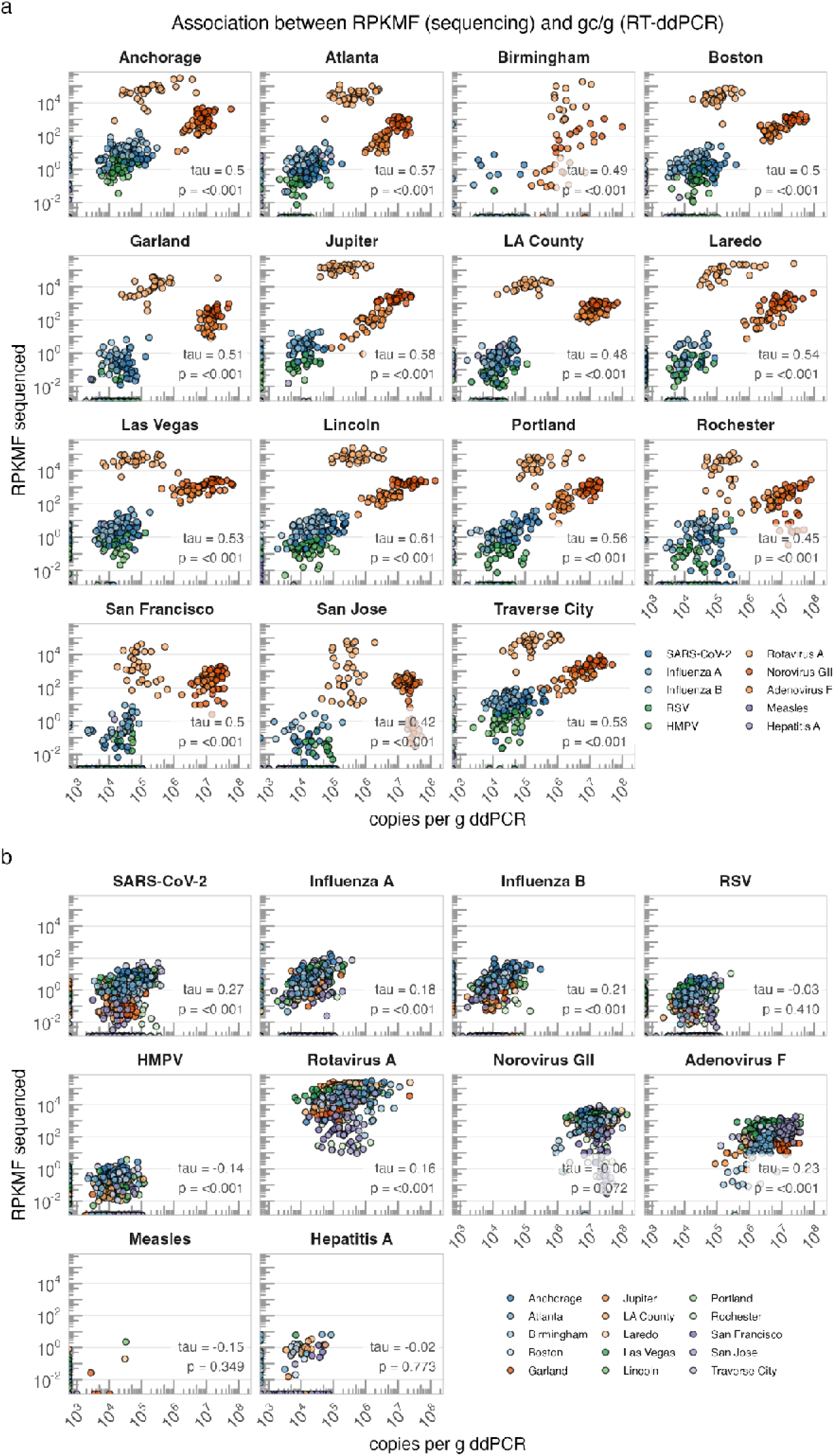
Relationship between relative abundance of viral targets via sequencing (RPKMF) and abundance in gc/g by RT-ddPCR for **a)** each location across all viral targets with color representing different targets, and **b)** each viral target across all sampling locations with color representing locations.

### Measles sensitivity and genotyping

Measles was detected 7 times throughout the sampling period via RT-ddPCR: 4 times in Garland, TX; 1 time in Lincoln, NE; 1 time each in Laredo, TX, and San Jose, CA (Fig 7, Table S7). Of these 7 detections, 3 coincided with sequencing detections. The RT-ddPCR assay used is inclusive of both wild strains of measles genotypes D8 and B3, but does not distinguish between the two^4^. Measles was therefore chosen as an example to further explore the classification of sequencing data by genotype, using the same pipeline described above. Measles was detected via sequencing in 29 out of 443 samples across the sampling period, in 7 out of 15 locations. All alignments to measles obtained >96.6% average read identity, representing the percentage of read bases matching the reference genome, but read counts were typically low, with 18/29 samples containing <10 measles reads (median = 8, range = 1-1,070, median percent coverage = 2.7%, range = 0.8-96.8%) (Table S7). Of these samples, 86% (24/29) produced reads that predominantly mapped to genotype D8. In three samples, measles reads were attributed to genotype A, corresponding to the measles vaccine. Two other genotypes were identified in wastewater samples: D4, which was circulating globally during the study period and was identified in a sample from a location with multiple B8 detection in other samples (1 sample; 2 reads, 99.3% identity, 0.90% coverage breadth), and C1, a genotype that is not known to be currently circulating (1 sample; 4 reads, 99.27% identity, 1.72% coverage breadth) (Fig 7, Table S7). There was only one sample in which reads were attributable to multiple genotypes, both D8 and A.

**Figure 7.**
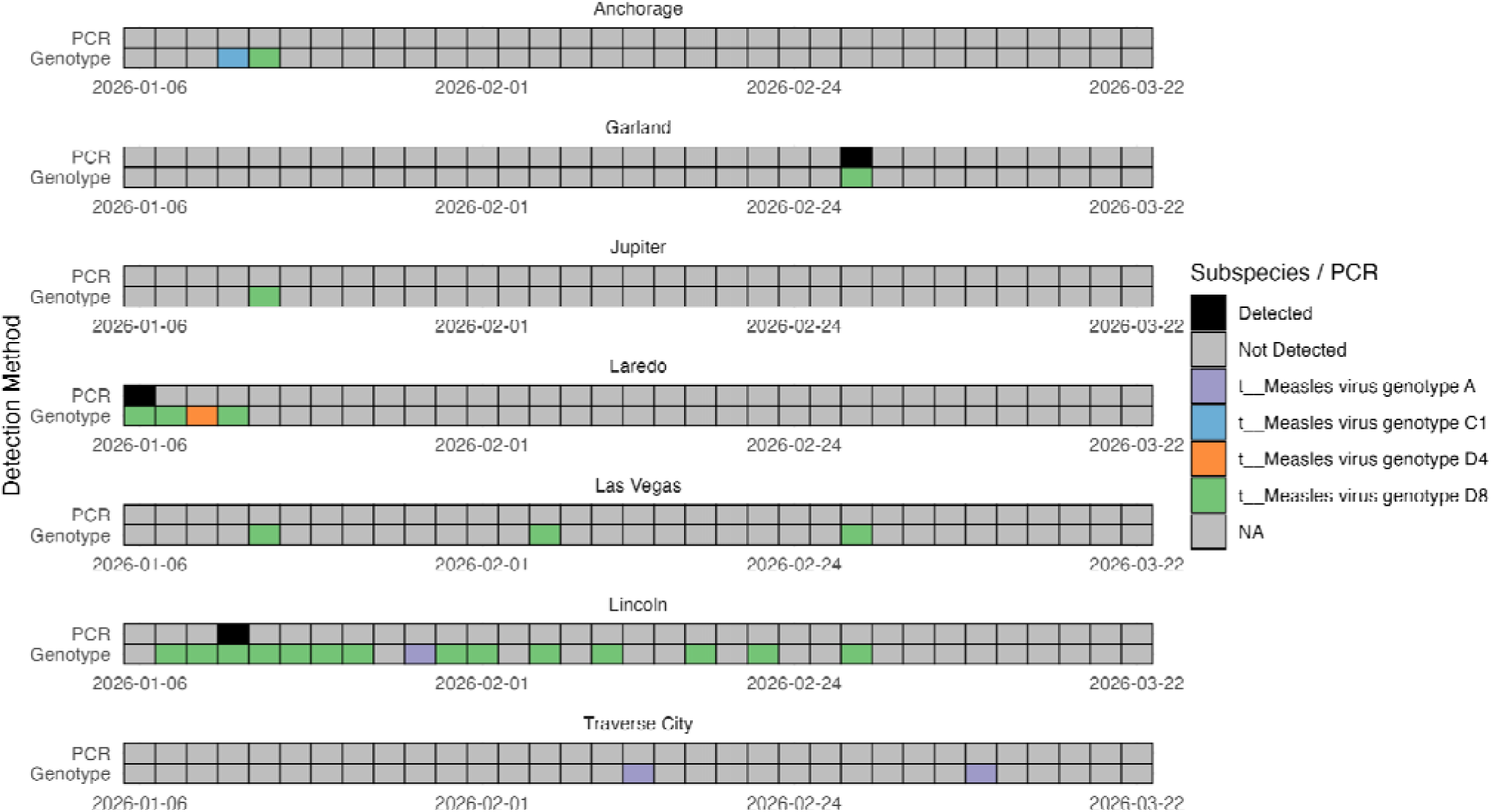
Heatmap showing detections of measles by both RT-ddPCR (top row) and sequencing (bottom row) by WWTP. Color represents the genotype of measles associated with the reads from each sample. Please note that different reads from the sample from Lincoln on 2/6/2026 were attributed to both genotypes D8 and A.

In one sample from Lincoln, 1,070 reads were attributed to genotype D8, covering 96.8% of the reference genome, demonstrating the potential to sequence complete pathogen genomes from wastewater metagenomes (Table S7). Three detections were attributed to genotype A (two in Traverse City, one in Lincoln), which is attributed to the measles vaccine. Overall, hybrid capture sequencing was more sensitive than RT-ddPCR for measles detection, and enabled genotype differentiation, which is critical for public health interpretation of circulating strains.

### Manual and semi-automated inspection of reads assigned to pathogens of concern

Reads assigned to pathogens of concern, including H5N1 Influenza A virus, Mpox virus, Marburg virus, poliovirus, swine vesicular disease, and avian paramyxovirus 1, using the bioinformatics pipeline (Table S8) were further investigated using additional direct validation steps as described in the methods. Using the polio direct validation pipeline, we determined that none of the read pairs classified as poliovirus represented a true detection of poliovirus; they were closer matches to non-polio enterovirus C or to conserved regions of the enterovirus C genome. Avian paramyxovirus 1 read pairs were determined not to span the fusion protein cleavage site that defines them as velogenic^41^. All H5N1 read pairs were confirmed as described above. All mpox virus read pairs were confirmed to align to mpox and other *Poxviridae*. Swine vesicular disease read pairs were determined to be better matches to other enterovirus B species rather than swine vesicular disease. Lastly, the Marburg virus read pairs did not align to any known circulating Marburg virus, but were sequences present only in a synthetic Marburg virus genome used in research of antibody-mediated virus neutralization. Specifically, the reads were aligned against synthetic orthomarburgvirus (GenBank MK271062.1, 100% nucleotide match, 100% query cover), reference orthomarburgvirus (RefSeq NC_024781.1, 0% nucleotide match, 0% query cover) and a recent wild type orthomarburgvirus (GenBank PV700537.1, 0% nucleotide match, 0% query cover). Subsequent detections of Marburg virus in these samples all displayed the same pattern, aligning only to the synthetic orthomarburgvirus vector. The metagenomic reads are only aligned to the synthetic orthomarburgvirus, being present in a region of the genome entirely absent from both the reference genome and the wild type sequences (Fig. S10).

## Discussion

In this study, we demonstrate that hybrid capture metagenomic sequencing **effectively enriches wastewater samples for sequencing of human viruses, enables sensitive, high-quality detection of rare viral targets, and produces results that are in strong alignment with quantitative RT-ddPCR measurements of key targets across diverse U.S. sewersheds.** Previous studies have demonstrated success in sequencing human viral targets in wastewater. We extend this work to evaluate the performance of hybrid capture sequencing for wastewater in 15 locations across the United States with a probe panel targeting a large suite of human viruses. Our findings for respiratory viruses are consistent with limited prior studies using shotgun and hybrid capture metagenomics, and we extend this comparison to a broader diversity of enteric and vaccine-preventable viruses. Importantly, we were able to detect nearly full reference genomes for many targets, including measles.

Hybrid capture metagenomic sequencing resulted in effective enrichment of a broad set of viruses in wastewater samples. While the percent of reads that were viral varied across WWTPs, viral reads were enriched by orders of magnitude above what has been reportedly obtained using shotgun metagenomics. One third of WWTPs (5/15) had a median percent viral reads higher than 25%, demonstrating consistently high performance at these WWTPs. The Twist Comprehensive Viral Research Panel used in this study targets 3,153 viruses, enabling broad capture of viral diversity in wastewater samples. We detected 1,334 different viral species across 443 samples, indicating diverse viral capture.

To assess whether hybrid capture sequencing can serve as a semi-quantitative alternative to PCR methods, we compared sequencing-derived reads per kilobase of genome per million filtered reads (RPKMF) with RT-ddPCR-measured genome copies per gram of dry-weight solids across 11 targets. RT-ddPCR concentrations were a significant predictor of sequencing-derived RPKMF across all 15 sites, with much of the variance in the LMM attributable to differences across viral targets and WWTP. Associations were consistent across sites whether the extracted matrix was influent or sludge, with a median Kendall’s tau of 0.51 across sites. Agreement in detection was strongest for consistently abundant enteric viruses. Rotavirus A, norovirus GII, and adenovirus F all achieved >99% positive agreement. The results reflect previous limited studies on respiratory viruses, showing similar agreement between signals from shotgun metagenomics and other probe-based hybridization approaches^16,24,27^.

While RT-ddPCR was more sensitive than sequencing for most viral targets, positivity rates were similar across methods even for rarer targets. In one case (measles), sequencing produced more detections than RT-ddPCR. This could be due to a variety of factors. For example, hybrid capture probes have a high tolerance to sequence divergence while RT-ddPCR assays need to be highly specific. Further, using sequencing, a sample can be scored positive for a virus if any portion of its genome is present whereas for RT-ddPCR, a sample is only scored as positive if the selected portion of its genome that is targeted by the assay is present and successfully amplified. The latter may be particularly important when concentrations of the target are low.

An important advantage of hybrid capture sequencing over PCR approaches is the ability to identify subtypes or characteristics of viral genomes. Targeted PCR assays amplify a single, predefined genomic region and are less robust to single nucleotide variants and other evolutionary changes in a genome. This makes these assays vulnerable to false negatives when circulating variants accumulate mismatches at primer or probe binding sites, a challenge that requires continuous assay redesign, particularly for pathogens with frequent changes such as influenza and measles^8^. PCR assays typically target highly conserved regions of the viral genome, and multiple assays are needed in order to detect different sub-species or genotypes depending on which targets are important for public health goals. Hybrid capture probes theoretically tolerate up to 30% sequence divergence and interrogate multiple regions of each viral genome^42^, providing built-in resilience to variant evolution and reducing the operational burden of continuous assay maintenance. This promiscuity allows for the generation of data that can be used to identify subspecies and genotypes/strains with minimal, if any, changes in the laboratory methods.

Sequencing yielded substantial genome coverage for several priority pathogens, including SARS-CoV-2, influenza, and RSV (52.38%, 17.65%, 13% of detections attaining ≥50% coverage of the genome, respectively), which can enable downstream analyses such as subtyping and phylogenetic placement^43^ (Fig. S11). Previous studies have shown that this enables tracking of H5N1 influenza subtypes^44^. Additionally, reads distributed across the viral genome can be assembled into larger contigs or even near-complete genomes. We recently employed this *de novo* assembly approach to recover divergent fragments of the influenza A genome and link them to avian-associated lineages^43^. Success of subtyping for viruses depends on variability in the viral genome, depth of sequencing coverage, and the inclusion of circulating subtypes in databases. While approaches will need to be tailored to specific viral targets, this sequence-level resolution positions hybrid capture metagenomics to greatly broaden the questions addressable by WBE.

Recovery of duplicate reads – likely from single cDNA fragments that are sequenced multiple times – in a sequencing effort is expected to be due to over-sequencings. This can occur when input cDNA has low sequence diversity, suggesting that viral enrichment did not occur as intended. In the present study, we identified samples with high-quality viral sequences as those with a duplication rate <99%. Duplication rate ranged from a median 96% to 99% across sites; sequencing was halted partway through the study at Birmingham, which had >99% duplication rate in most samples and the lowest number of unique reads among all sites (median = 2.53*10^6^). There was no significant difference in duplication rate by sample matrix, however the number of viral reads per site did appear to differ by matrix, suggesting that influent samples may yield higher-quality data in terms of quantity of viral reads. Despite filtration and CRISPR-based rRNA depletion, prokaryotic rRNA still accounted for a median of 7.7% of reads, indicating that off-target nucleic acids remain a meaningful portion of the nucleic acids sequenced. Sequencing depth was also a key determinant of viral recovery, with both total viral reads and unique viral species scaling log-linearly with read pairs sequenced. These observations suggest that for hybrid capture WBE, sample matrix selection, upstream depletion of host and bacterial nucleic acids, and per-sample sequencing budget jointly govern the sensitivity of the assay.

Several limitations should be considered when interpreting these results. Probe-based hybridization was chosen for this work to enable sensitive and efficient detection of human viruses in wastewater; however, enriching the sample manipulates the material such that presence and quantity of different viruses may be over or under-estimated. When enriched, relative abundance of viral targets may be altered by the efficiency of viral capture. Viruses that are present in the sample but not targeted may not be sequenced. Another consideration for this study is that we used all mapped reads in viral classification, and did not implement a coverage cutoff or other metric to further filter detections. It is possible that implementing stricter thresholds may alter the overall estimated abundance of viruses in these samples and their relationship with other measurements such as those obtained using RT-ddPCR. Additionally, we acknowledge that this less stringent approach to coverage thresholding may result in false positive detections, particularly when reference genomes attain low breadth of coverage (percent of genome covered by reads). The identification of a rare measles genotype (C1) in a sample with only 4 reads and low coverage (1.72%) is an example of a subtype assignment that may be important to investigate further. However, manual inspection of reads is costly in terms of efforts and stricter thresholds may reduce sensitivity. In some cases, low breadth of coverage may result from mis-mapping of reads to genomic regions shared between related viruses, rather than true presence in the sample. Discerning true, low-abundance detections from spurious ones will require further, careful work that balances the risk of false positives with that of false negatives, the latter of which may have serious implications for public health response. Here, we present all results to demonstrate the potential of targeted viral sequencing to provide nuanced information about both common, higher coverage pathogens as well as rarer ones, rather than a definitive accounting of viral presence/absence over our time series. Finally, viral targets referred to in this study are exclusive of prokaryotic viruses (bacteriophages), which are likely to account for larger biomass than human viruses in these samples.

It is important to note that the matrix-type comparisons in this study compare two different sample sources rather than partitioning behavior of viruses between liquids and solids. Metagenomic sequencing was performed on the liquid fraction of both sample types. Sludge-derived samples were centrifuged prior to processing to remove excess solids, whereas influent samples were processed without removal of solids. Future studies that include direct comparisons of sludge and liquid-derived samples from the same sites would also provide evidence for the best sample types for hybrid capture sequencing.

In summary, this study illustrates that hybrid capture sequencing can be scaled to generate information on presence and quantity of human viruses in wastewater from WWTPs across the United States, each with unique characteristics and population coverage, to infer information on infectious diseases circulating in the contributing populations. A remaining challenge is to understand how to best prioritize public health actionable data within the large amounts of information obtained with this approach, and how to move beyond tracking presence and abundance of targets over time. We anticipate that the data can be used to identify emerging viral infections and outbreaks, for situational awareness, and various response activities as is done with PCR-based data now. The scientific community should work closely with public health officials and clinicians to understand how to prioritize and communicate outcomes from these data and explore how subtyping and characterization of viral genomes may best serve the needs of the community.

## Data availability

Deduplicated sequencing reads are available through the NCBI under BioProject PRJNA1438722. Accessions are also listed in Table S1.

## Acknowledgements

We thank the wastewater treatment plant partners at A.K. Warren Water Resource Recovery Facility, City of Garland Rowlett Creek Wastewater Treatment Plant, City Of Rochester MN Water Reclamation Plant, Clark County Water Reclamation District Flamingo Water Resource Center, Deer Island Treatment Plant, John M. Asplund Water Pollution Control Facility, Loxahatchee River Environmental Control District, Portland Water District East End Wastewater Treatment Facility, RM Clayton Water Reclamation Center, San Jose-Santa Clara Regional Wastewater Facility, Southeast San Francisco Wastewater Treatment Plant, Theresa Street Water Resource Recovery Facility, Traverse City Regional Waste Water Treatment Plant, Village Creek Water Reclamation Facility, and Zacate Creek Wastewater Treatment Plant for assistance with sample collection. We thank the local and state public health department partners in Alabama, Alaska, California, Georgia, Florida, Maine, Massachusetts, Michigan, Minnesota, Nebraska, Nevada, and Texas for their support of this work.

