## Supplementary Information for "Prospective metagenomic sequencing of wastewater across the United States yields robust viral enrichment and concordance with digital PCR measurements"

*Measles assay*

Until March 2, 2026, the measles assay was used with only “probe 1”, as indicated in table S1. The methods and assay validation using probe 1 were previously published^1^. The measles assay using probe 1, as described in Boehm et al., was designed to detect both B3 and D8 measles genotypes^1^. The D8 genotype is the one that is mainly circulating in the U.S. at the time of this work, although imported cases of B3 from abroad have been identified. A B3 variant circulating outside the U.S. has a mutation (a single-nucleotide polymorphism) in the portion of the sequence where probe 1 binds (Canadian examples include accession numbers PQ873016 through PQ873023 in NCBI, accessed May 1, 2026). Therefore, we designed a second probe (probe 2) to capture that sequence, as shown below. Both probes are run in the same channel (FAM).

| Oligonucleotide | Sequence (5’-3’) |
| --- | --- |
| Probe 1 | CATGATGATCCAAGTAGTAGTGA |
| Probe 2 | CATGATGATCCGAGTAGTAGTGA |
| Forward Primer | AGGATGAGGCGGACCARTACTT |
| Reverse Primer | CRATATCTGAGATTTCCTTGTTCTC |

Primers and probes used to detect wild-type measles.

1.	Boehm, A. B. *et al.* Pathogen nucleic acids data in wastewater solids from 147 treatment plants in the United States: 2024–2025. *Data Brief* **65**, 112503 (2026).

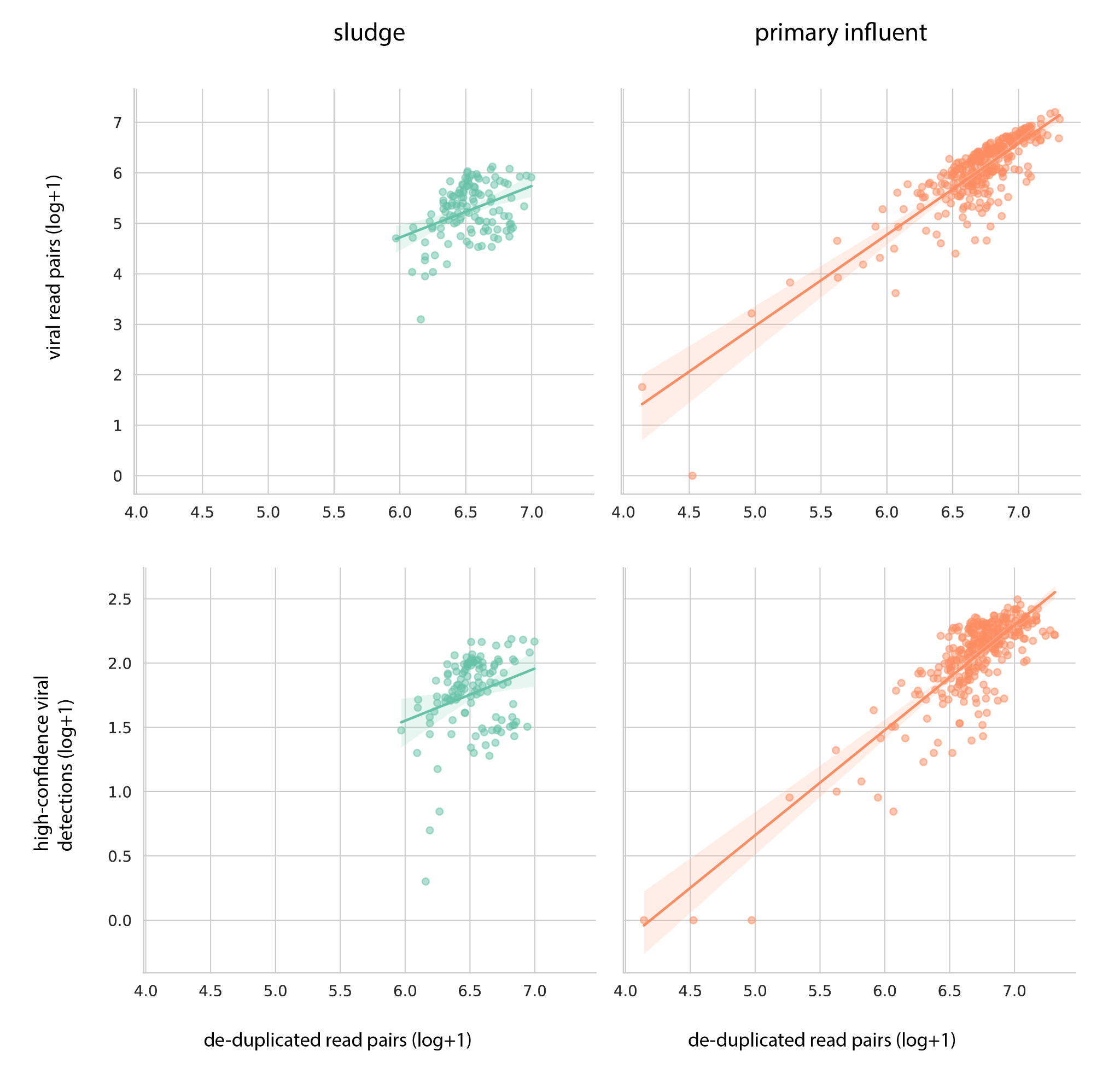

**Figure S1.** Relationships between sample matrix, sequencing effort, and viral detection. Top row: viral read counts as a function of total read pairs sequenced. Bottom row: high-confidence viral detections (≥50% coverage breadth, ≥1x coverage depth) as a function of total read pairs sequenced. All values are log-transformed.

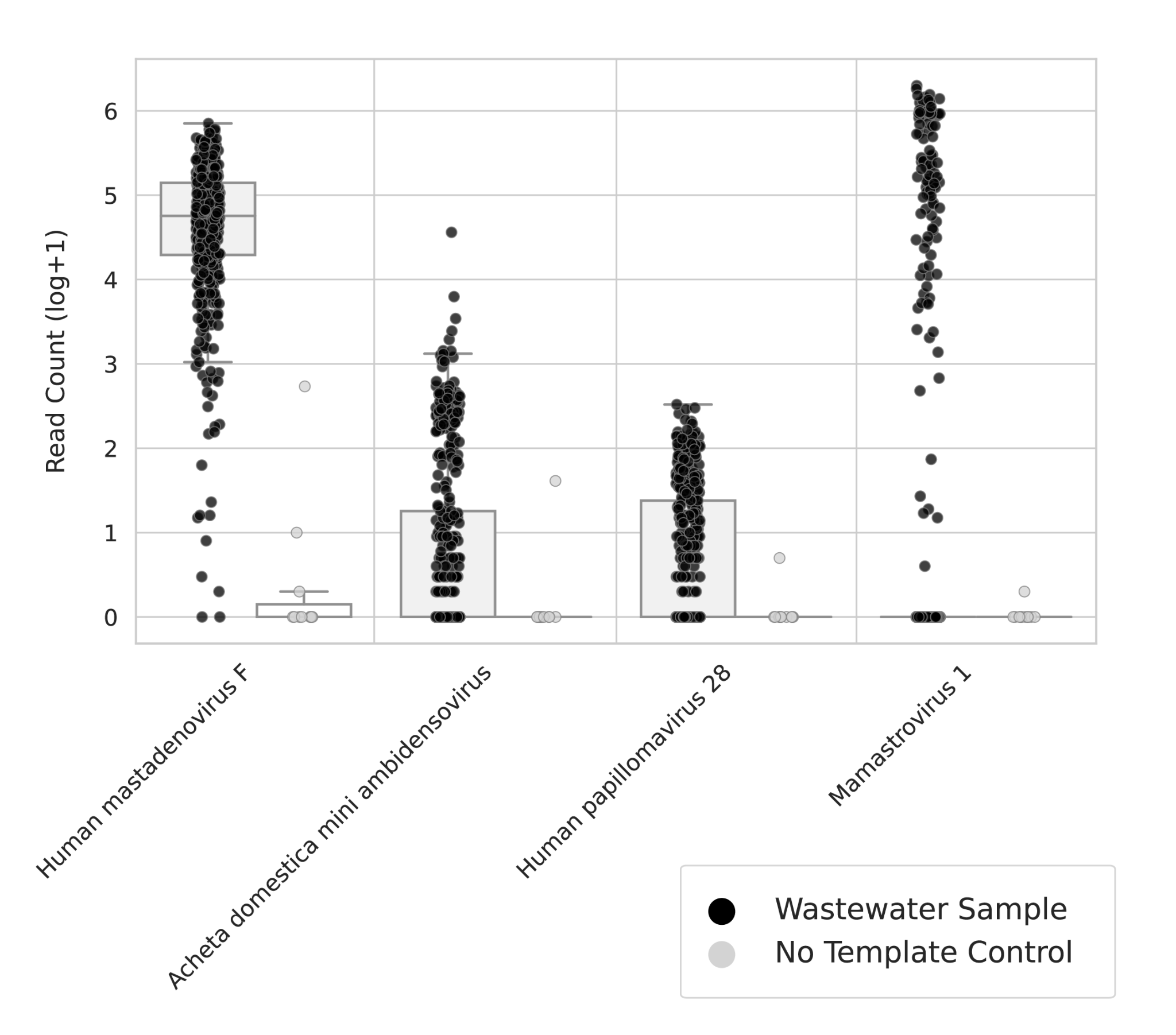

**Figure S2.** Log-transformed read counts for all viruses detected in no template controls. Read counts for these viruses are contrasted between no template controls and wastewater samples.

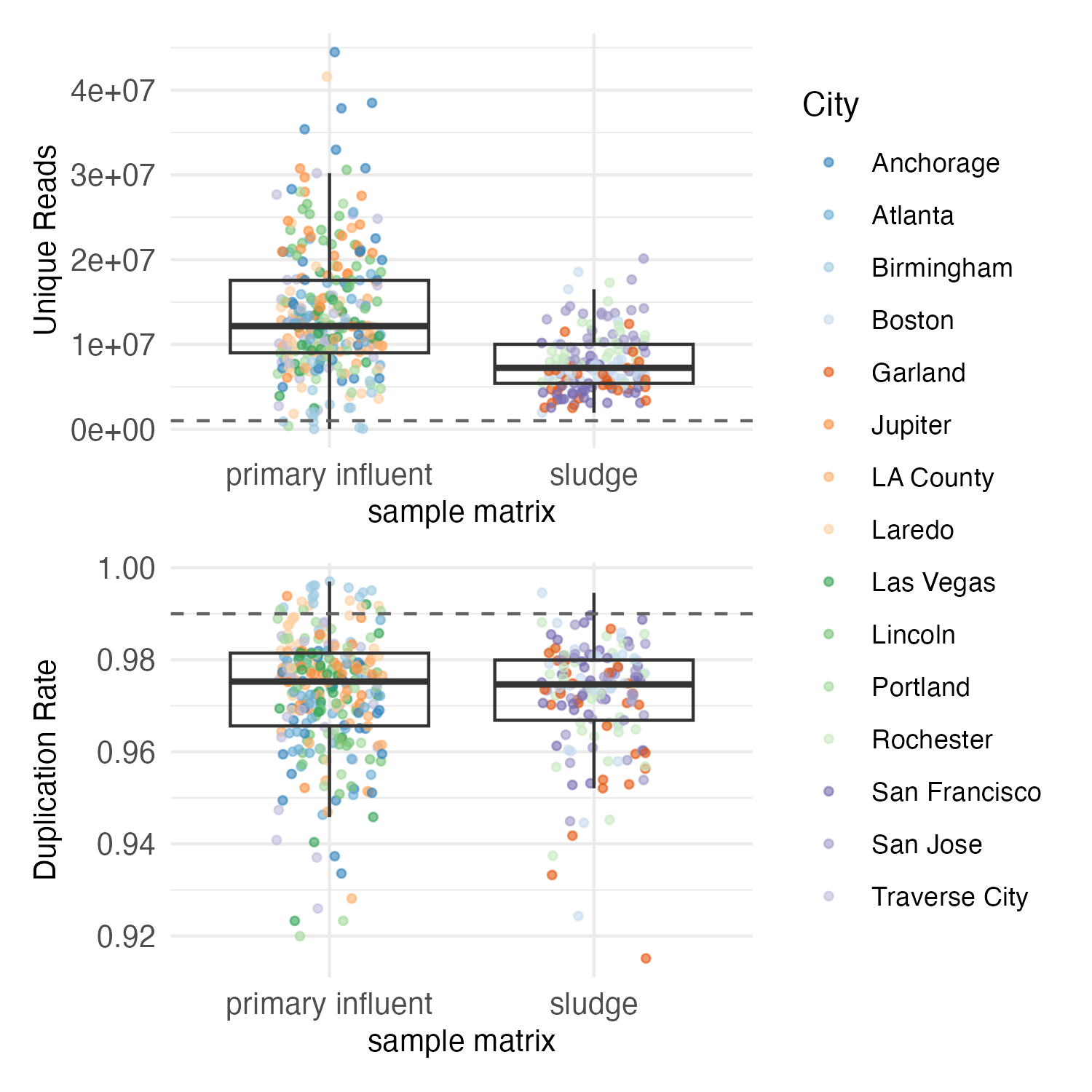

**Figure S3**. Sequencing quality outcomes in terms of unique reads post deduplication (top row) and duplication rate (bottom row). The dashed lines indicate the QC thresholds, which are 1,000,000 unique reads (top) and a 99% duplication rate (bottom).

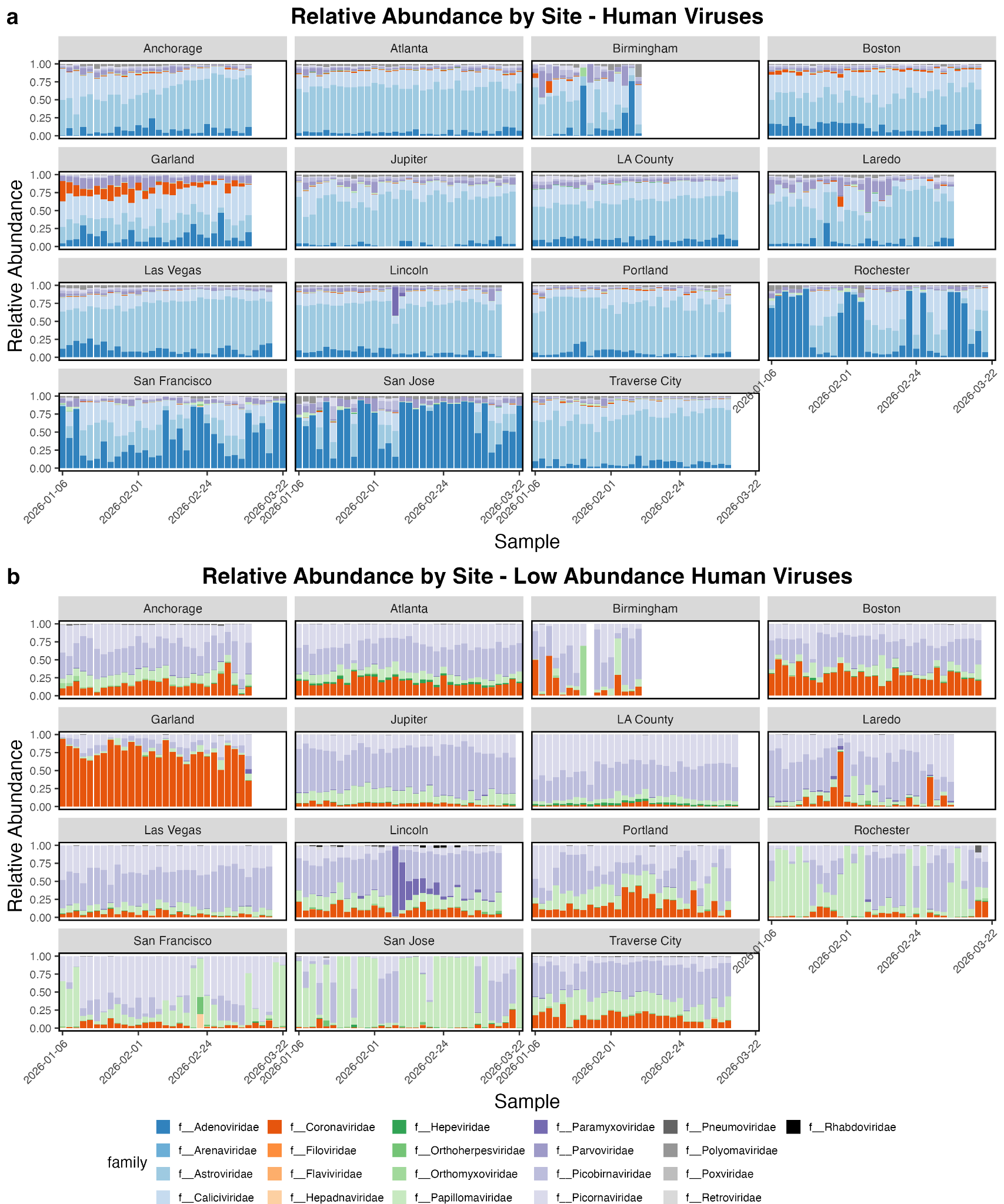

**Figure S4.** Relative abundances of viral families containing human pathogens. a) Relative abundances of all families containing viruses with human hosts. b) Relative abundances of families containing viruses with human hosts, with the six most abundant families overall removed to highlight lower-abundance targets.

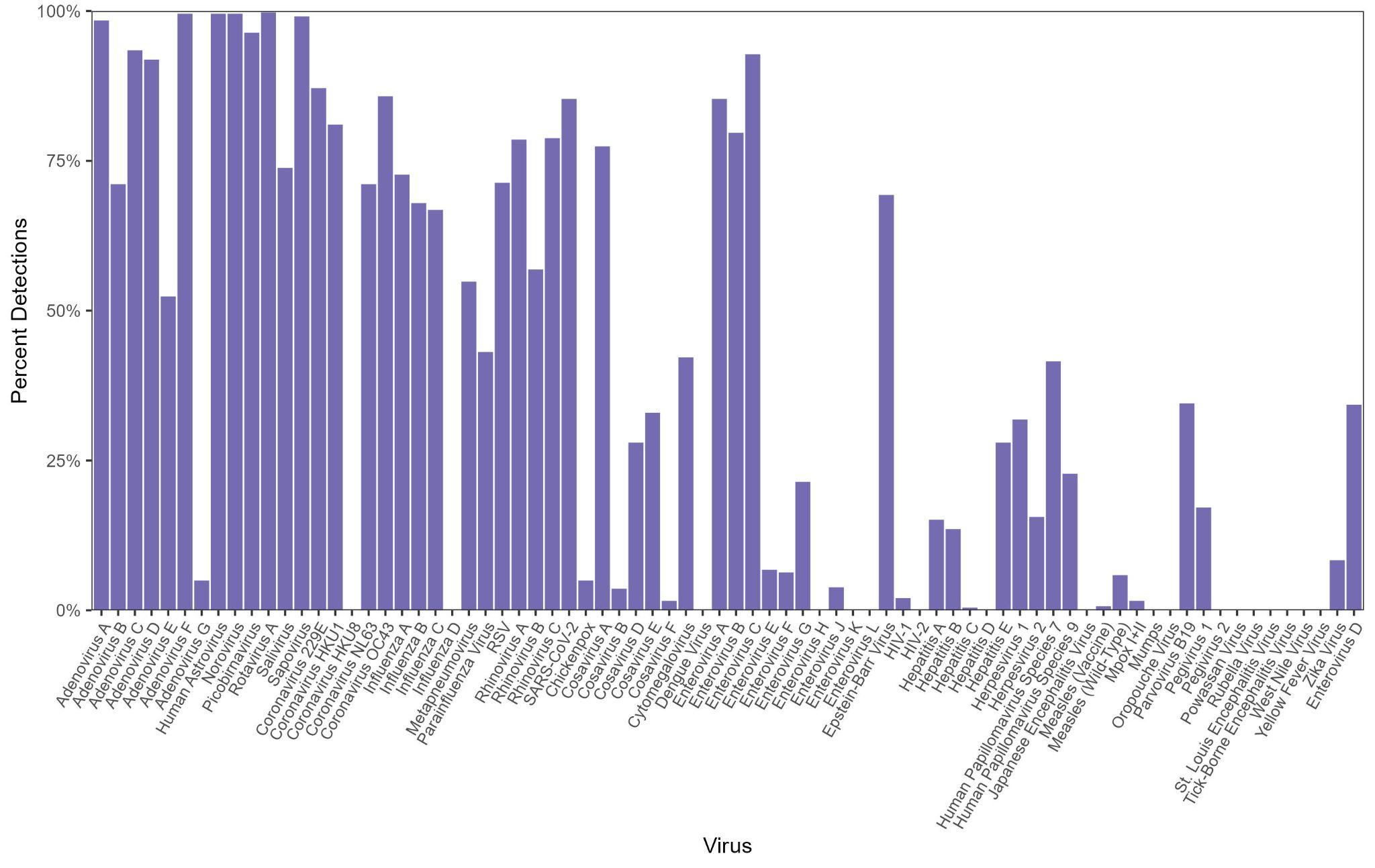

**Figure S5.** The percent of samples that each virus was identified in using EsViritu. This list only shows common pathogens as specified in Table S3 and does not include high-consequence diseases.

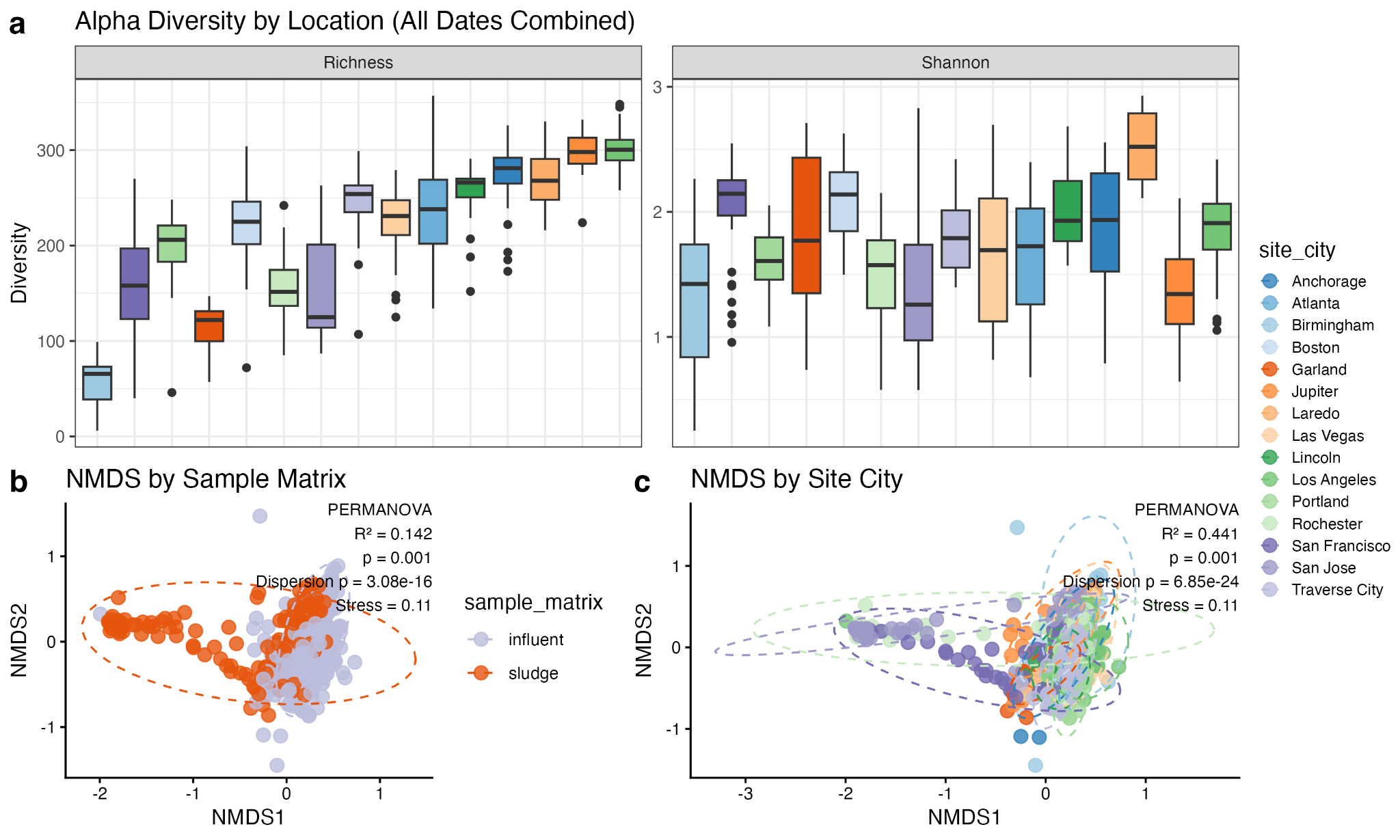

**Figure S6.** Alpha and beta diversity analyses. a) Boxplots showing the median and interquartile range (IQR) of species richness and Shannon diversity index for each WWTP, summarized across all sample dates. b) Non-metric Multidimensional Scaling (NMDS) of Bray-Curtis dissimilarity, colors and PERMANOVA results representing sample matrix impacts on sample divergence. c) Non-metric Multidimensional Scaling (NMDS) of Bray-Curtis dissimilarity, colors and PERMANOVA results representing WWTP impacts on sample divergence.

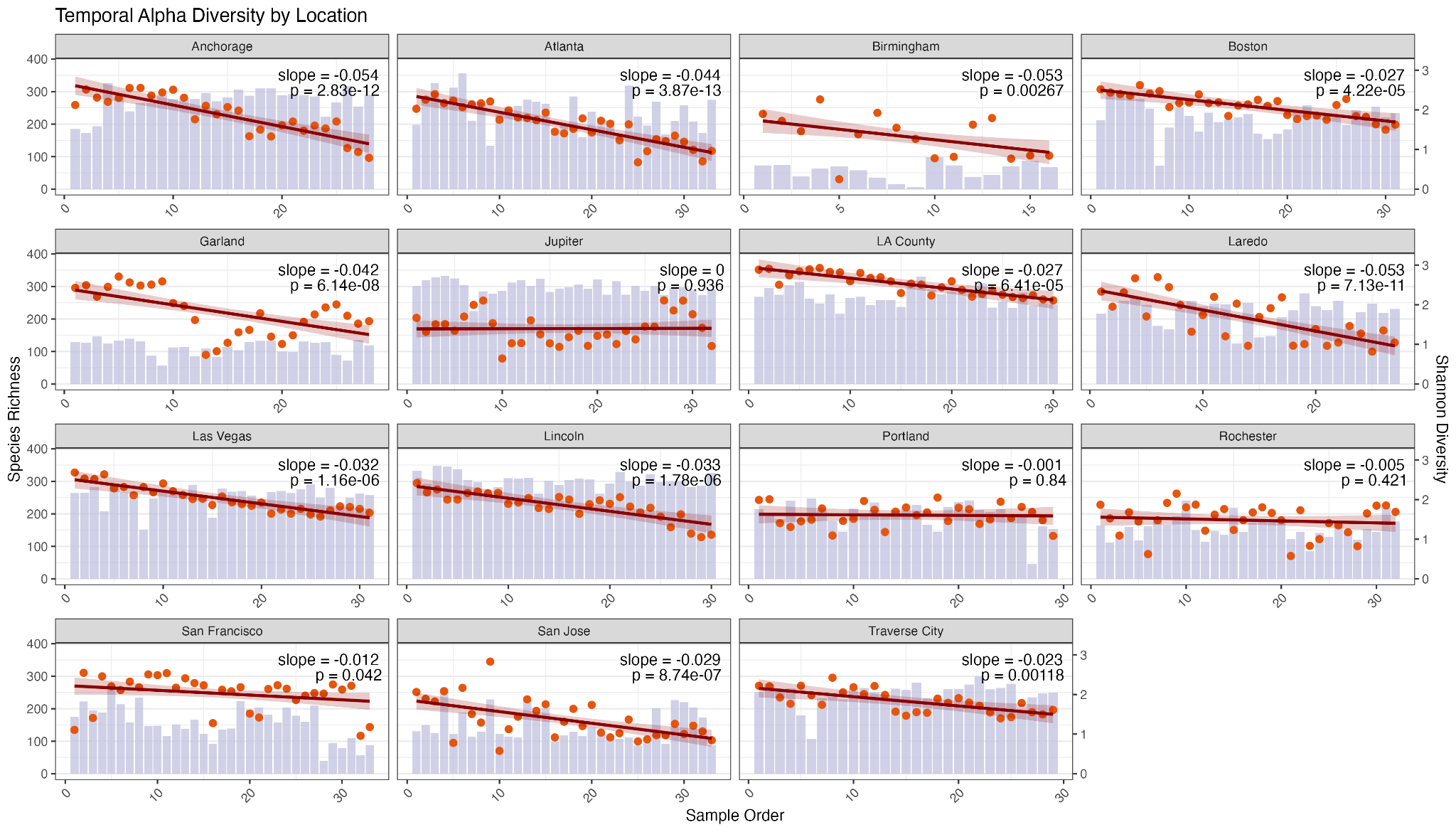

**Figure S7.** Species richness (purple) and Shannon diversity (orange) shown temporally for each WWTP location. Linear regressions are shown for each location with slope and R^2^ for each line.

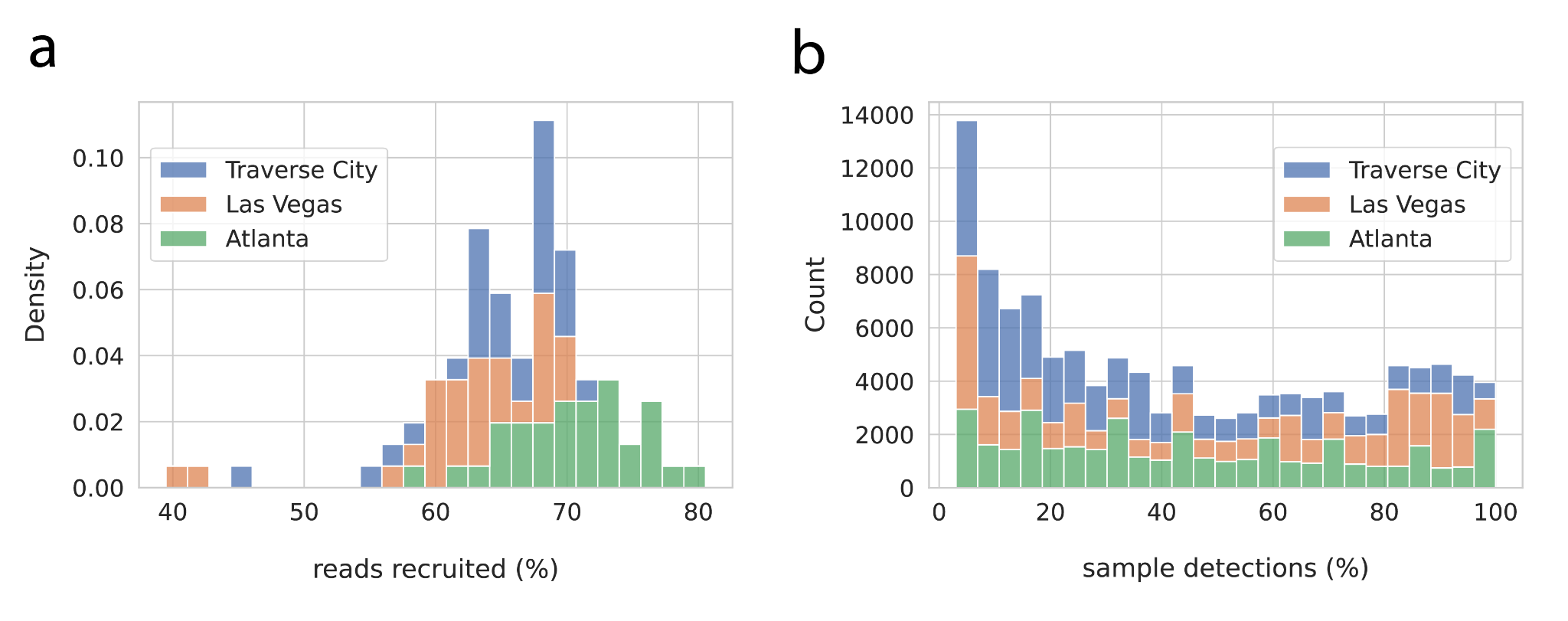

**Figure S8.** Features of *de* *novo* assemblies of ‘off-target’ sequencing reads (Materials and Methods). **a)** Percentage of filtered reads recruited by assembled fragments using loose identity thresholds. **b)** Frequency of detection for assembled fragments among samples at each site, using stringent identity and coverage breadth thresholds.

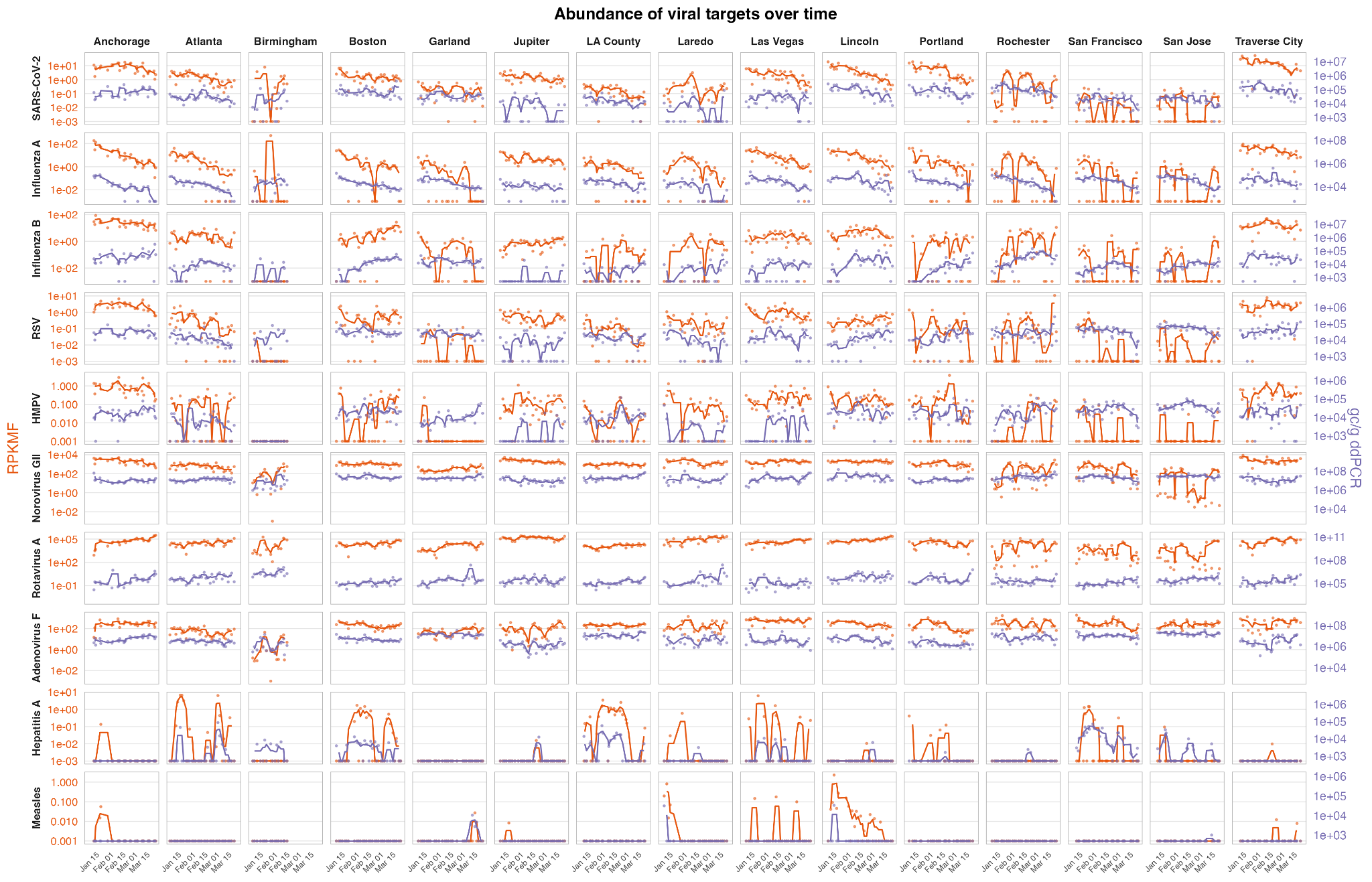

**Figure S9.** Time Series of RPKMF and gc/c by site and target. Points represent individual estimates of RPKMF (orange) and gc/g (purple) and lines represent a three sample rolling average.

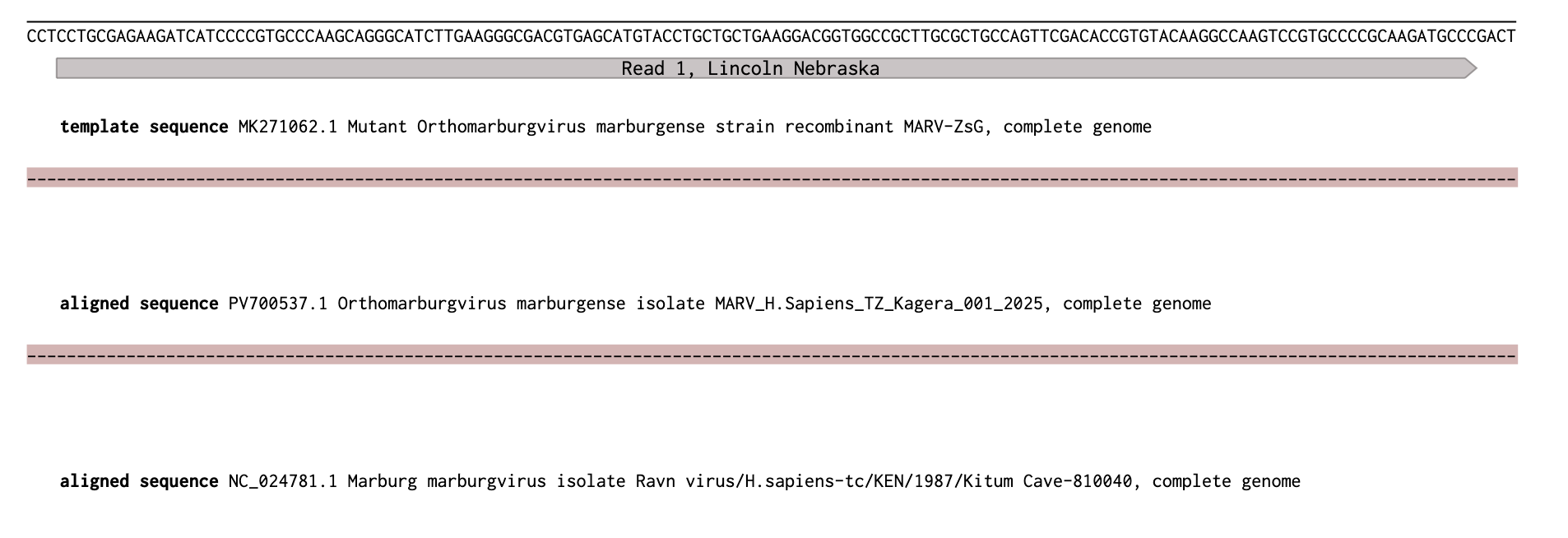

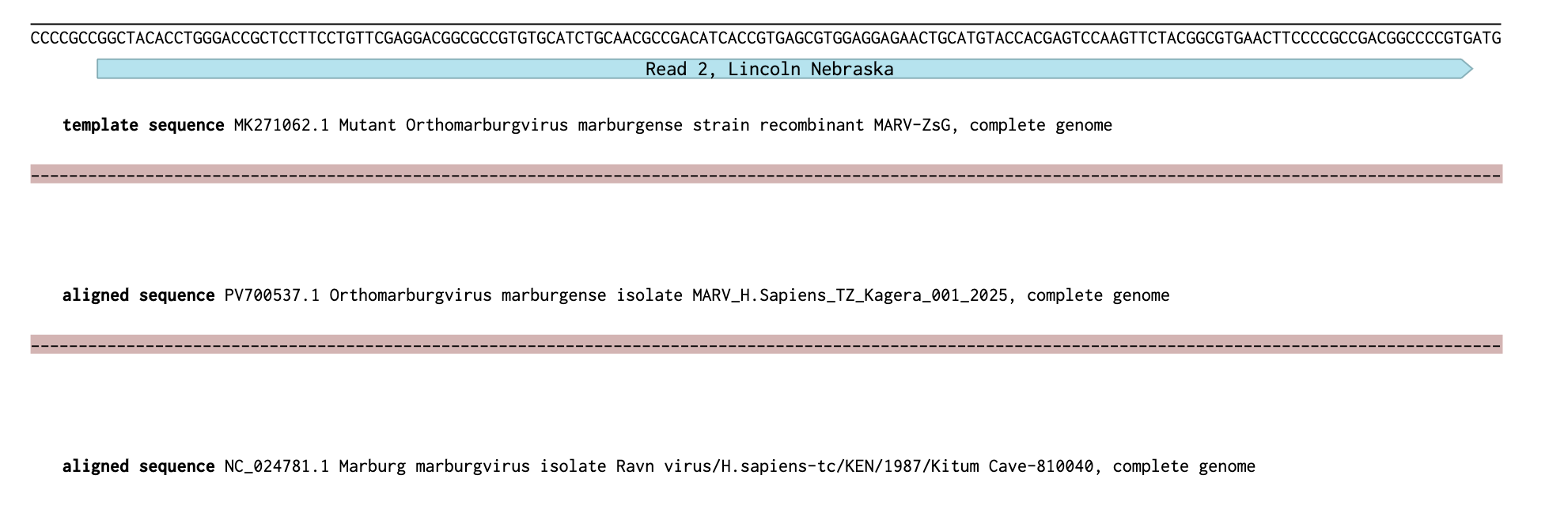

**Figure S10**. Alignment of Marburg virus reads from Lincoln, Nebraska, to synthetic genomes, circulating wild type genomes, and reference genomes. The entirely light red portions indicate the absence of those regions in the wild type and reference genomes. The top panel shows both reads, while the middle and bottom panel show zoomed in alignments to clearly demonstrate the missing regions.

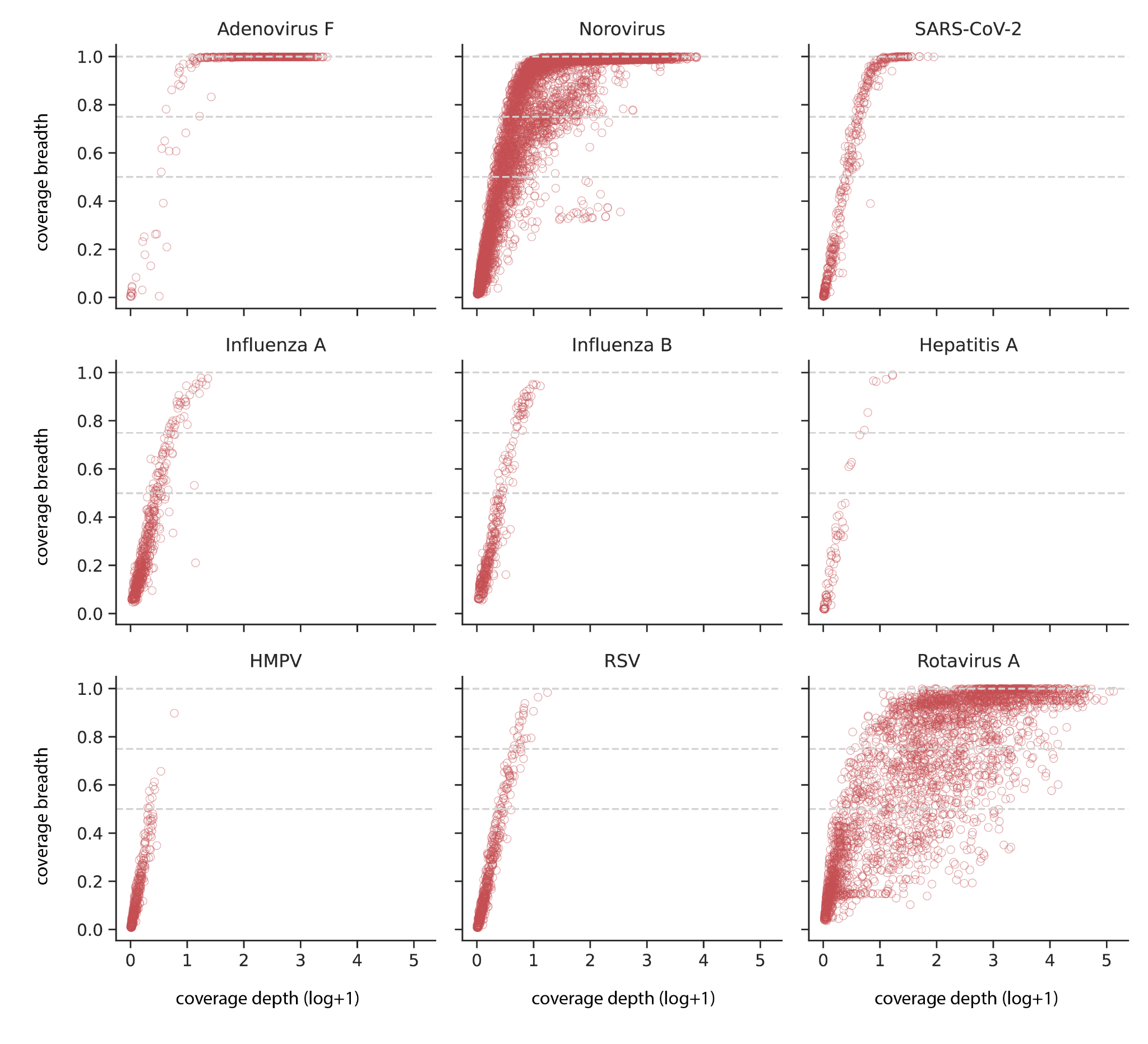

**Figure S11**. Sequencing coverage of priority pathogens. Coverage depth describes the mean sequencing depth across the reference genome; coverage breadth describes the proportion of total genome bases covered by reads. Each point represents a detection in a single wastewater sample.

**Heatmaps showing the relative abundance of common pathogens (Table S3) at each WWTP**
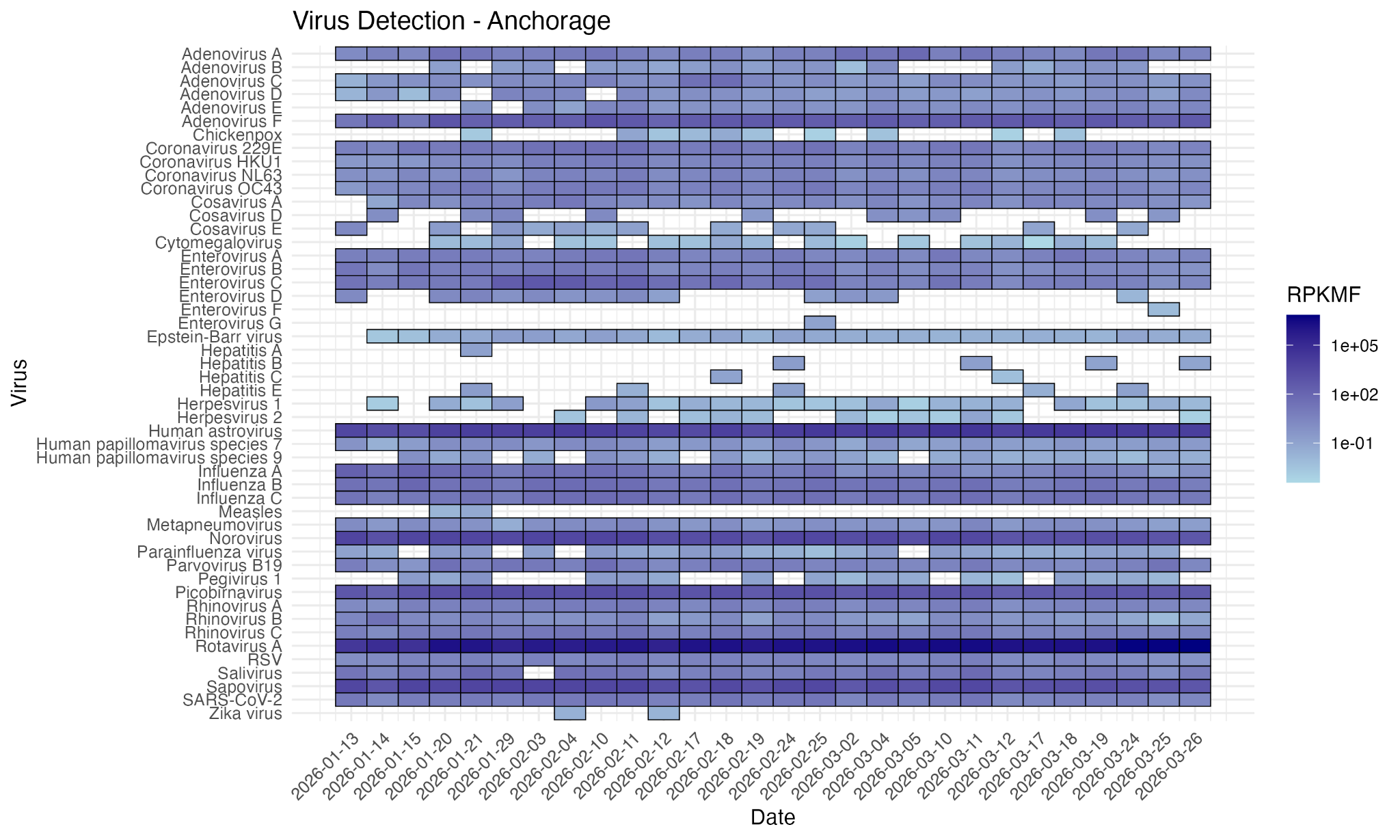

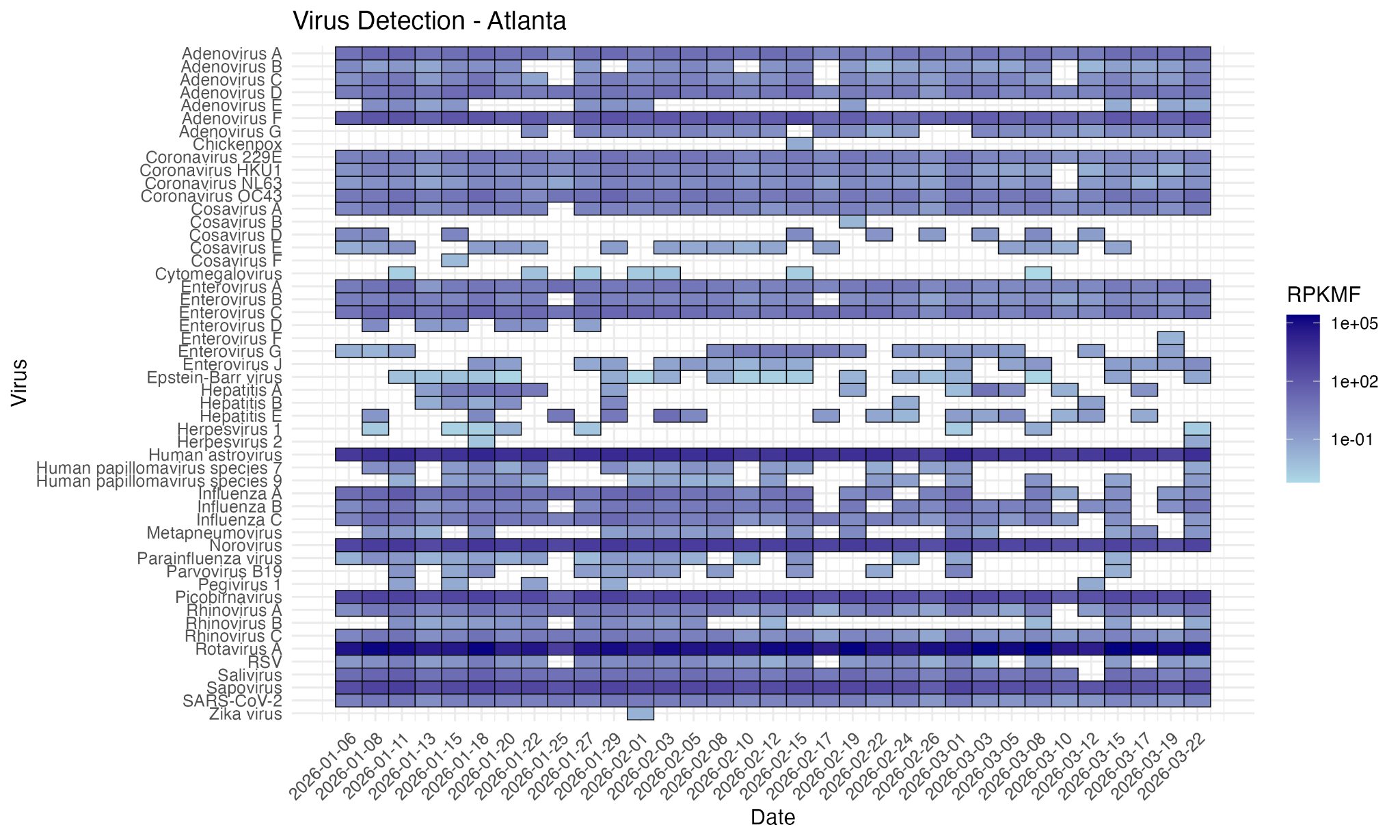

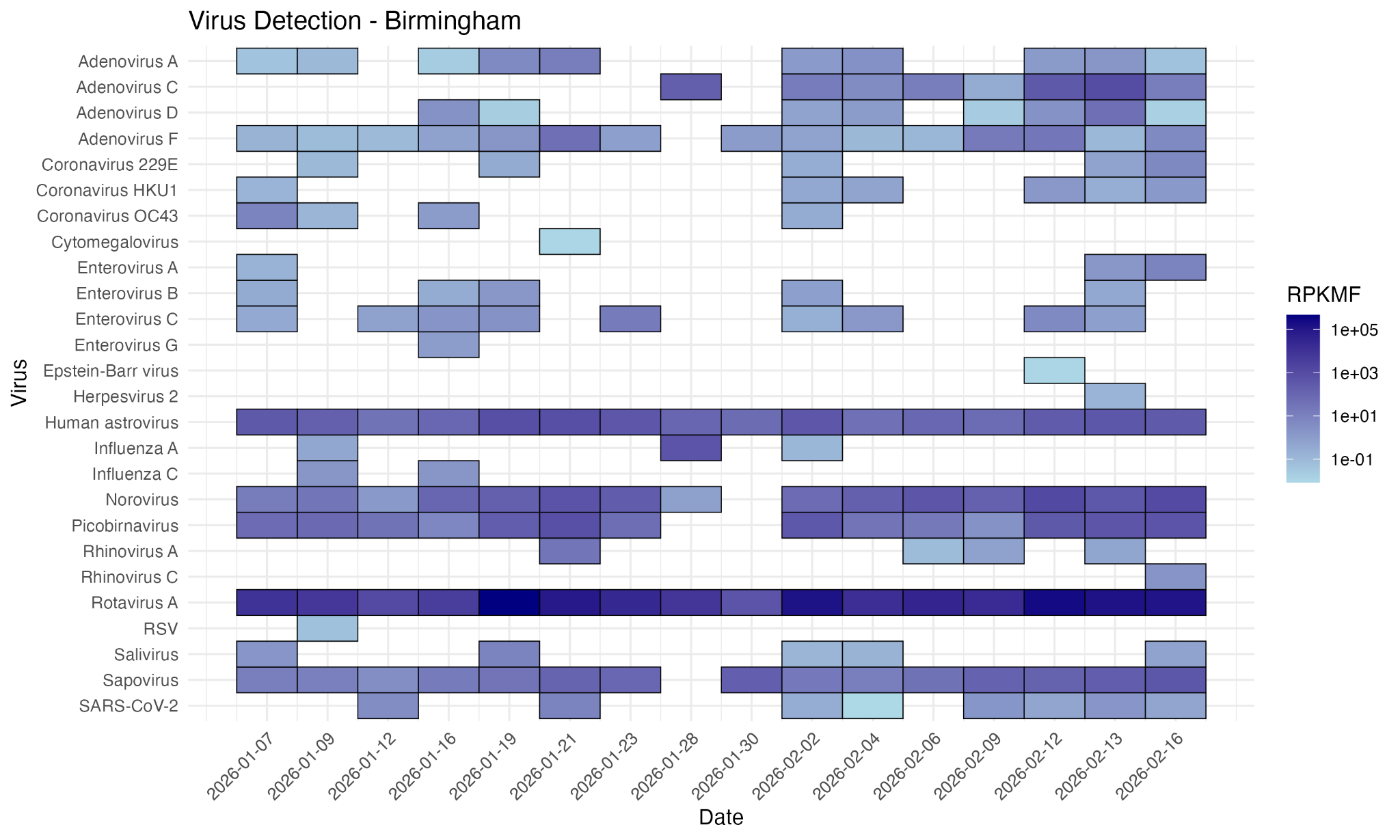

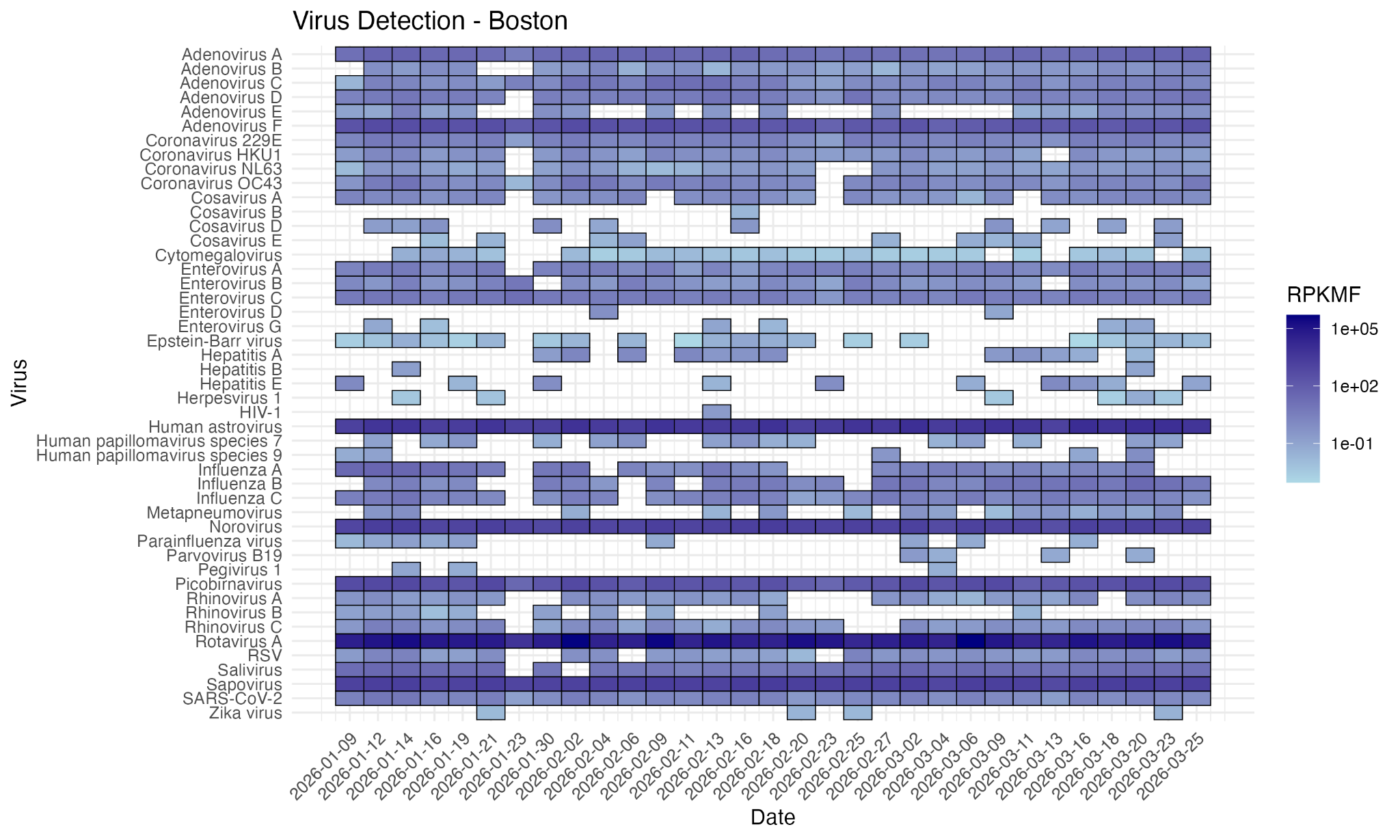

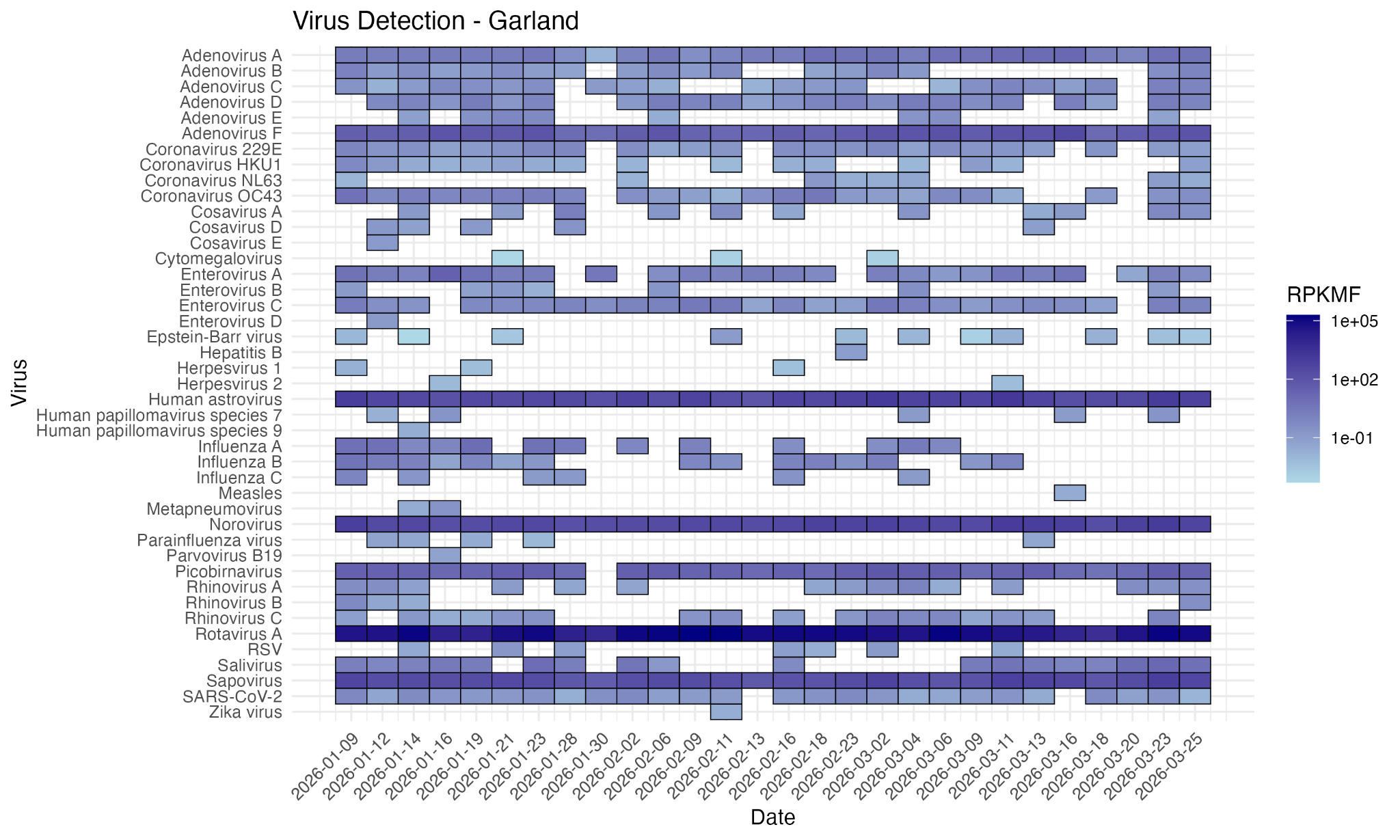

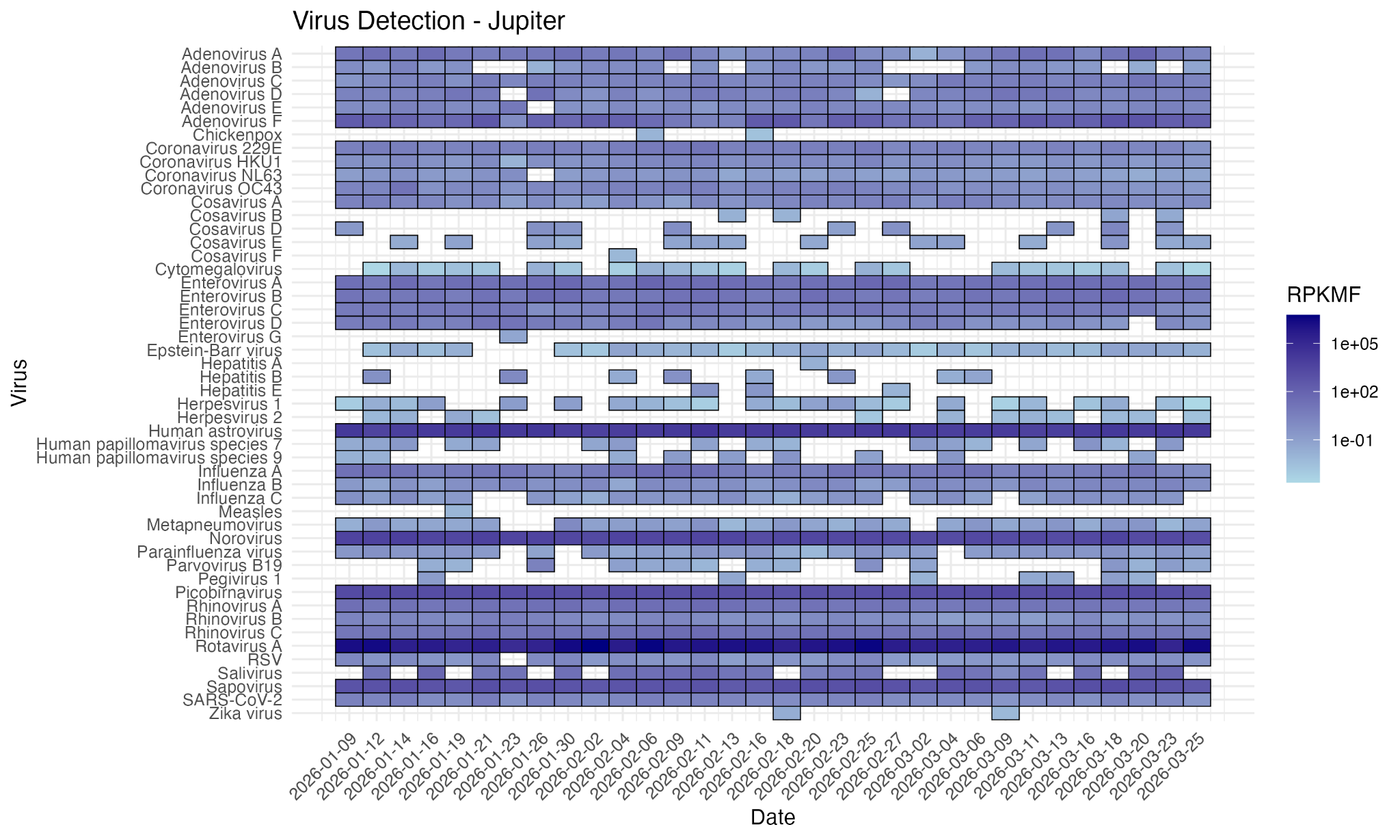

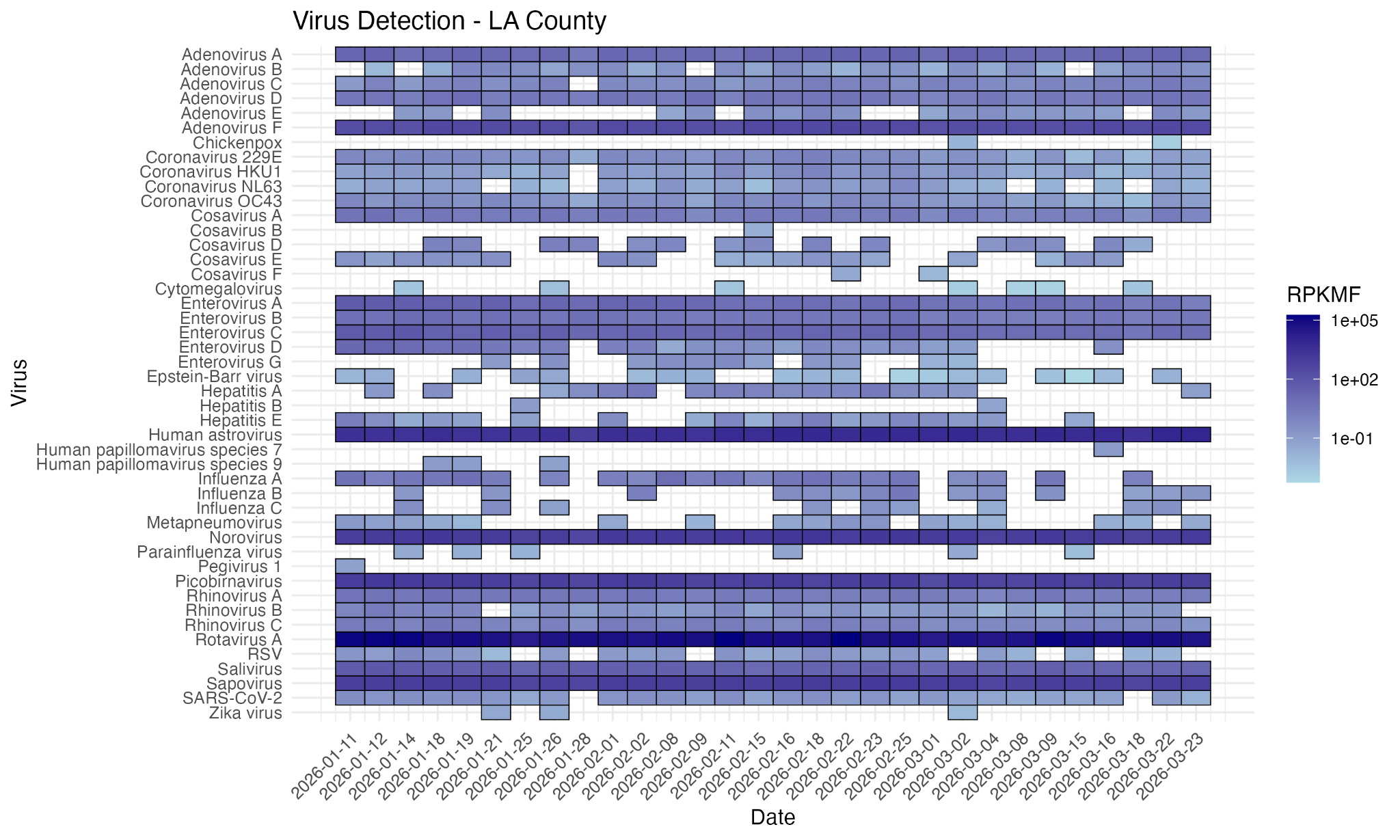

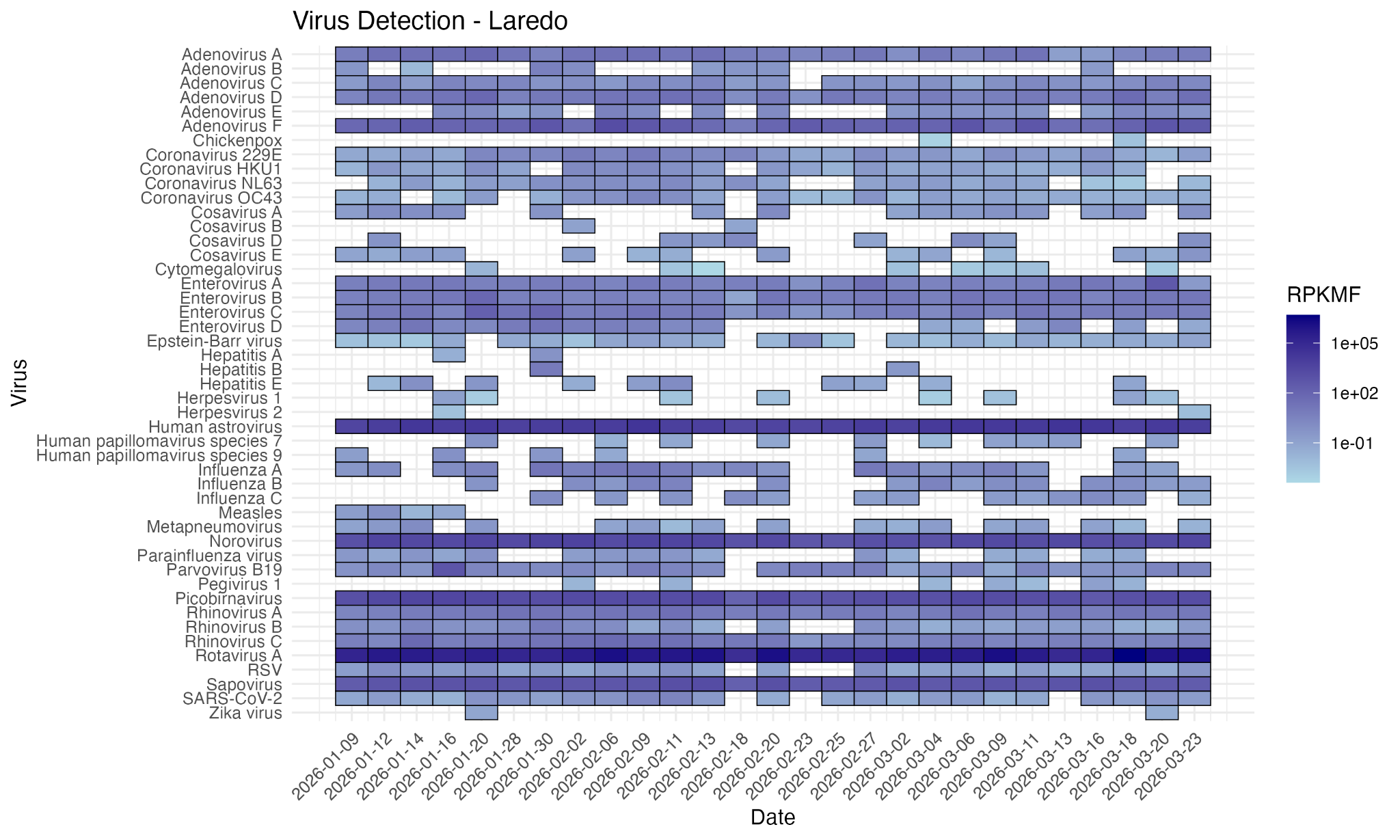

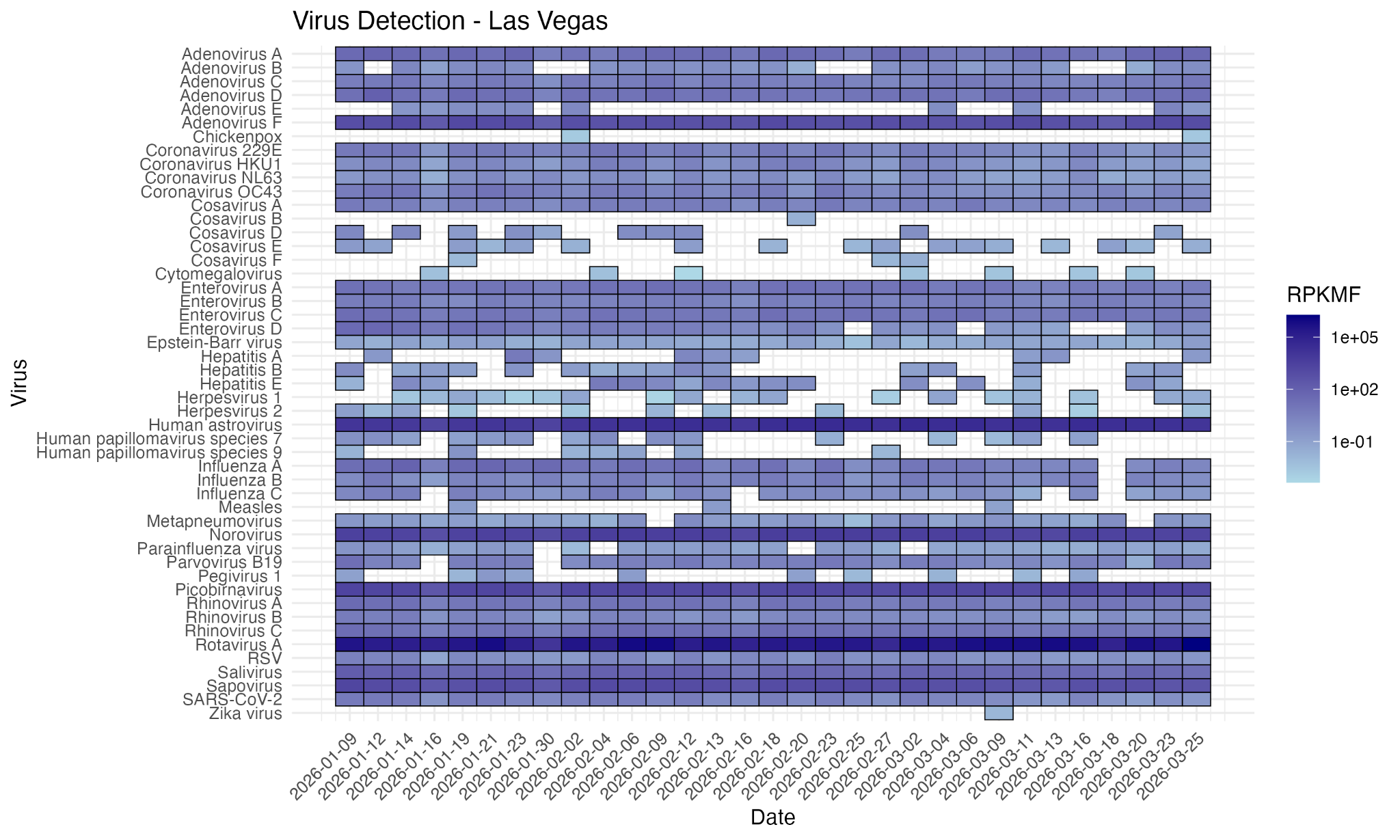

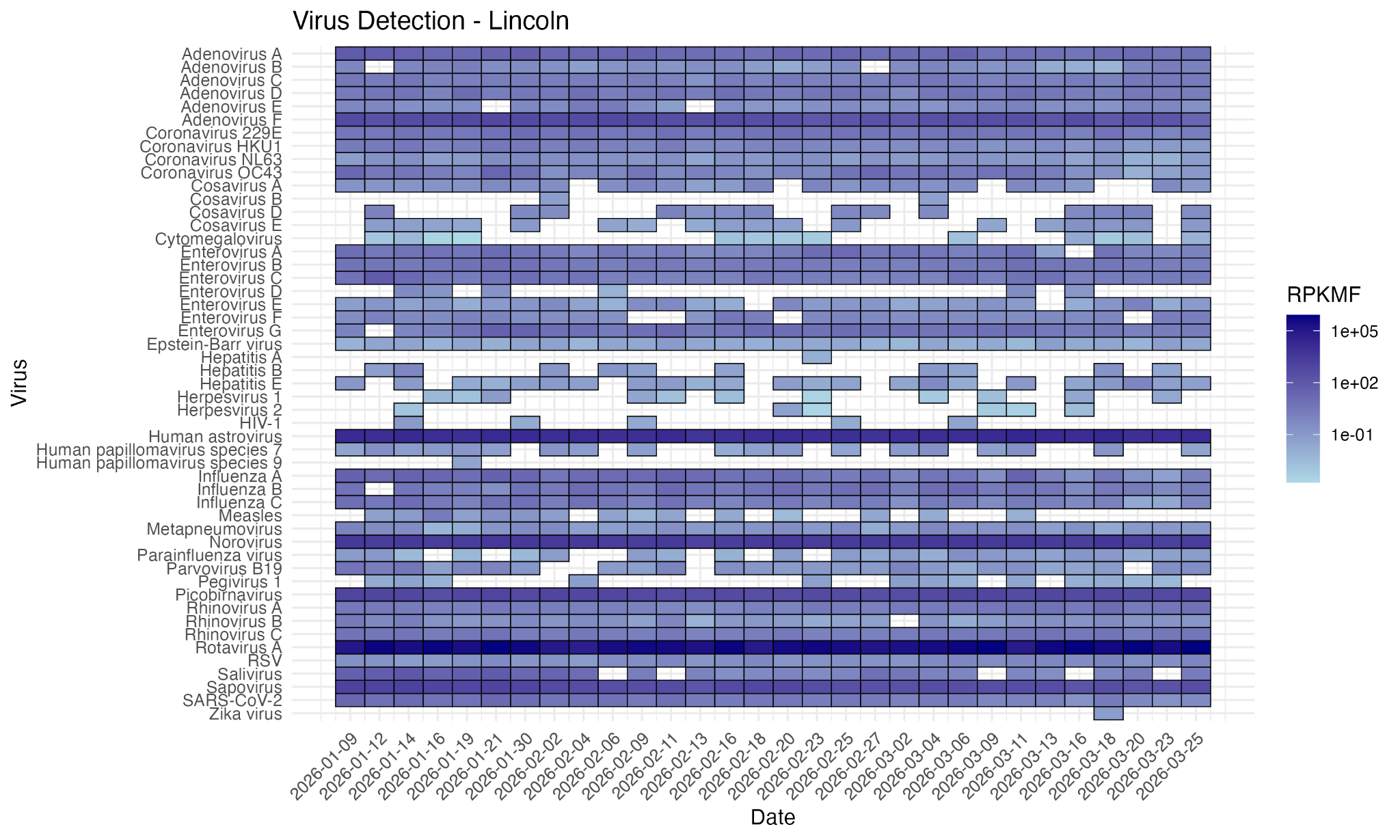

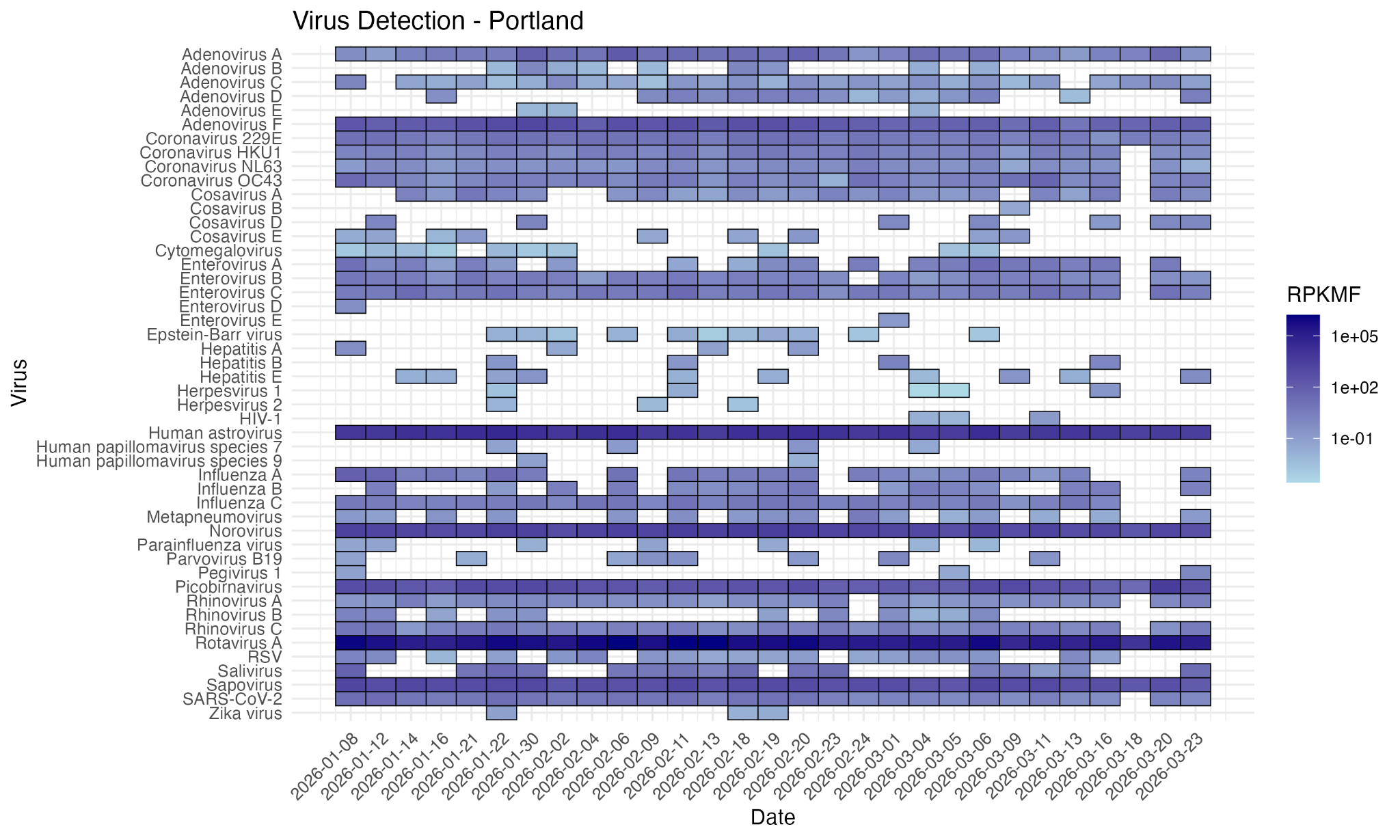

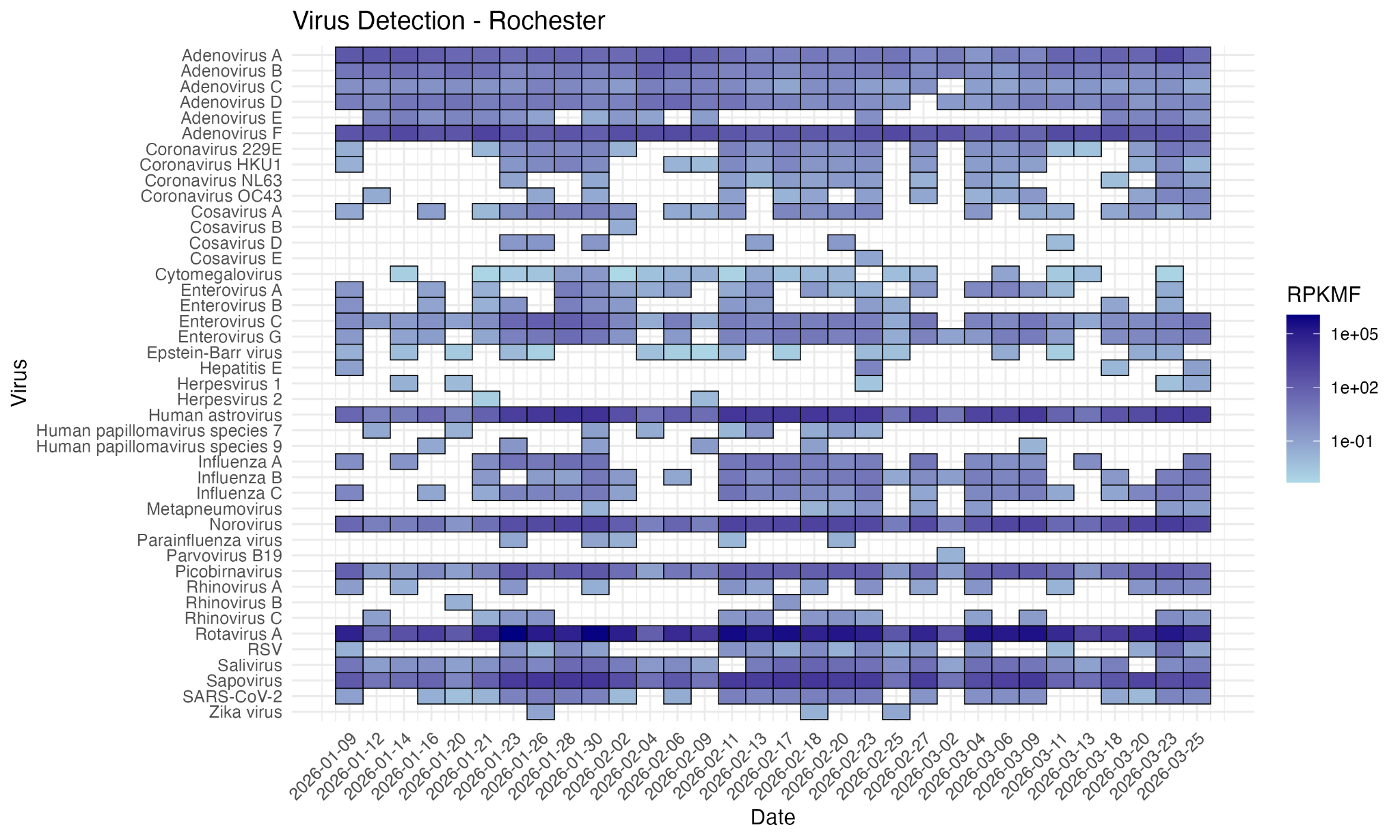

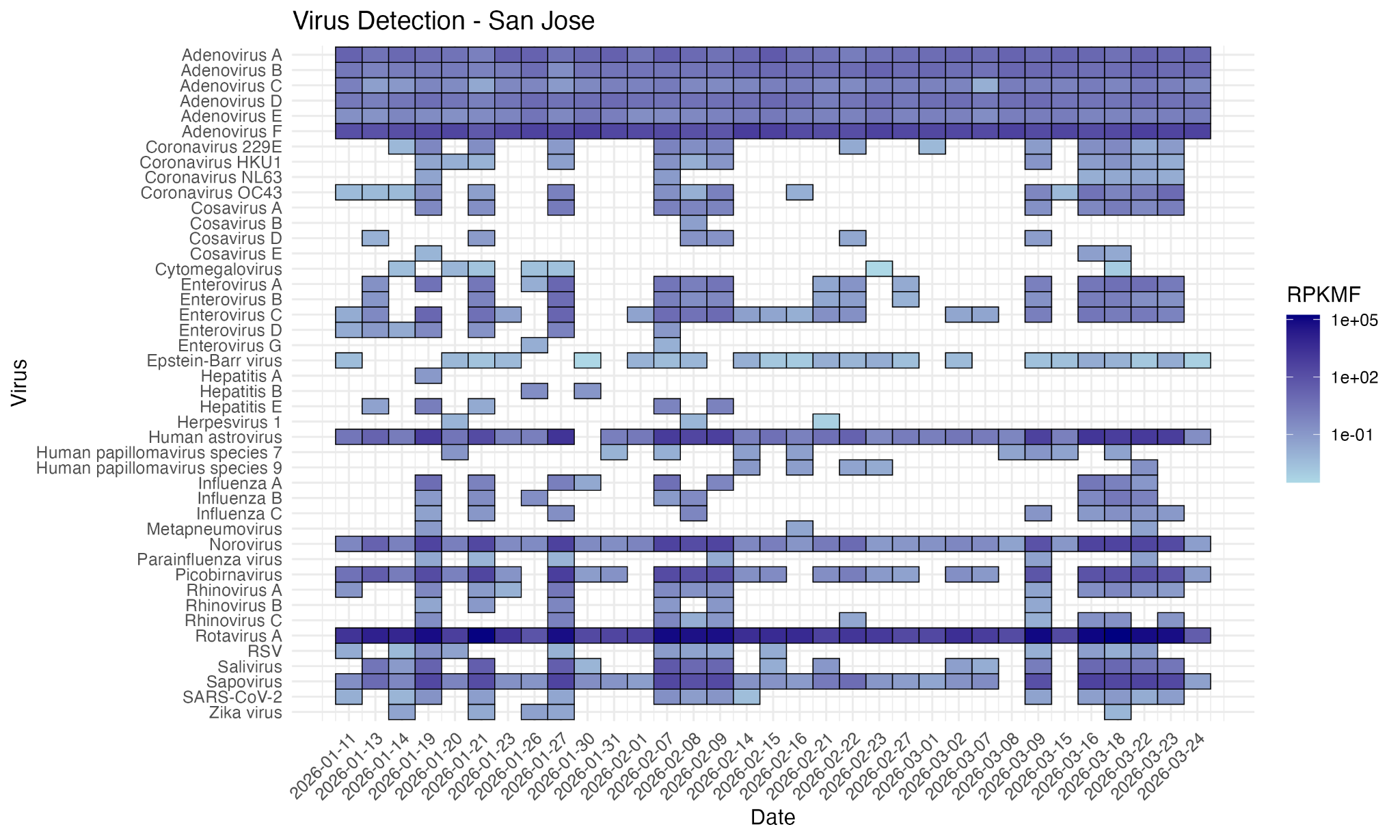

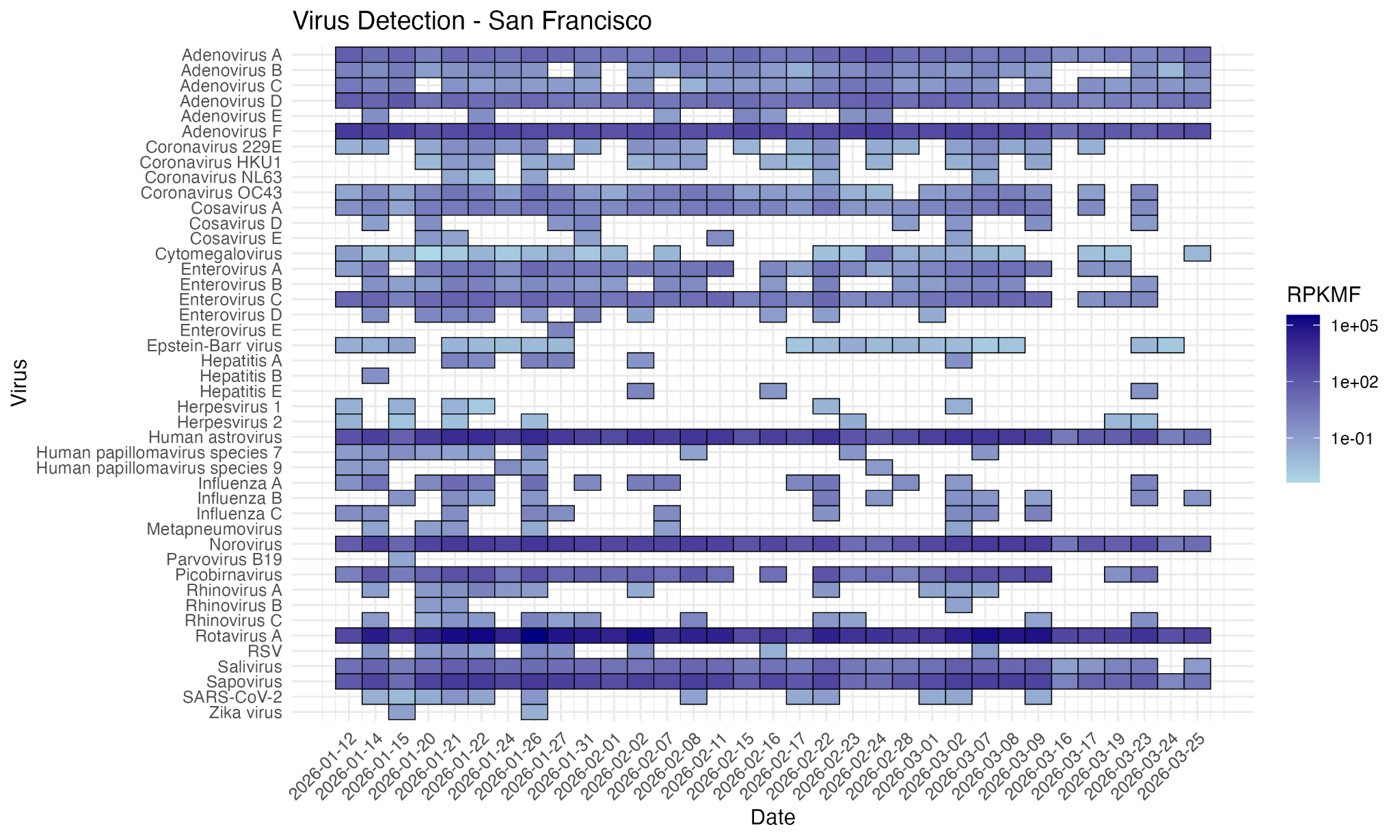
]
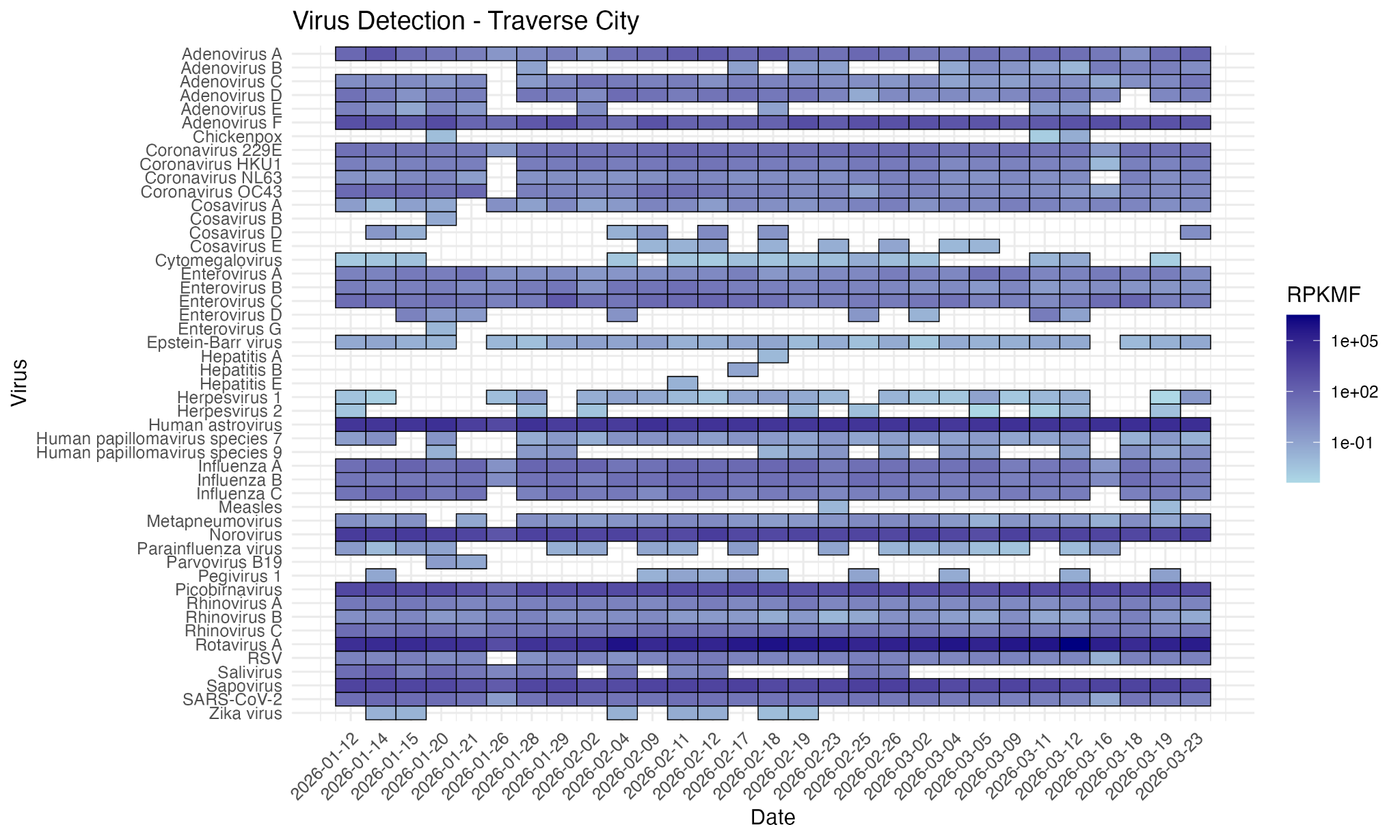
